# Mortality by cause of death in Brazil: effects of the COVID-19 pandemic and contribution to changes in life expectancy at birth

**DOI:** 10.1101/2023.02.13.23285842

**Authors:** Fernando Fernandes, Cássio M. Turra, Giovanny V. A. França, Marcia C. Castro

**Affiliations:** Demography Department, Cedeplar, Universidade Federal de Minas Gerais, Belo Horizonte, MG 31270-901, Brazil; Secretariat of Health Surveillance, Brazilian Ministry of Health, Setor de Rádio e Televisão Norte (SRTV) 701, Via W5 Norte, Edifício PO700. Brasília, Distrito Federal, 70719-040, Brazil; Department of Global Health and Population, Harvard TH Chan School of Public Health, Boston, MA 02115, USA

**Keywords:** Life expectancy at birth, COVID-19 mortality, time series decomposition, life expectancy decomposition, indirect effects of COVID-19 mortality.

## Abstract

We investigate the consequences of the COVID-19 pandemic on other underlying causes of death in Brazil in 2020 and 2021. We estimate monthly age-standardized mortality rates for 2010-2021 and decompose those time series into three additive components: trend, seasonality, and remainder. Given the long-term trend and historical seasonal fluctuations, we assume that any impact from the pandemic will be left on the remainder. We also decompose the contributions of COVID-19 deaths (direct effect) and those from other causes (indirect effects) to the annual change in life expectancy at birth (𝑒_0_) from 2017 to 2021. Broadly, the remainder mirrors the trajectory of pandemic waves. The impact of the COVID-19 pandemic on other causes of death was not limited to increases but also decreases. The direct effects of the pandemic reduced 𝑒_0_ by 1.89 years between 2019 and 2020 and 1.77 between 2020 and 2021. Indirect effects increased 𝑒_0_ by 0.44 between 2019 and 2020 and had virtually no impact on 𝑒_0_ between 2020 and 2021. Whether trajectories in mortality rates and annual gains in 𝑒_0_ will quickly return to pre-pandemic levels depends on governmental actions to mitigate the consequences of the COVID-19 pandemic.

## Introduction

The death toll of the COVID-19 pandemic is overwhelming. In Brazil, the country with the second-highest worldwide deaths, COVID-19 directly claimed about 638,000 lives between January 1, 2020, and December 31, 2022 – 9.5% of the global deaths (Brasil, 2022a, 2022b; World Health Organization, 2023). Since 1990, the Brazilian federal and local governments have implemented and expanded a unified, decentralized, and free public health care system (Sistema Único de Saúde – SUS) (Castro et al., 2019). Nevertheless, the absence of federal government action led to unequal and uncoordinated state and local government responses to the COVID-19 pandemic (Ventura et al., 2021). Combined with socioeconomic inequalities, these responses resulted in an unequal social and spatial burden of COVID-19 deaths (Castro, Kim, et al., 2021; Rocha et al., 2021).

However, the mortality burden of the COVID-19 pandemic is not limited to the deaths directly attributed to COVID-19 but also the indirect effects on other underlying causes of death (Castro et al., 2022). Indirect effects could manifest in different and contradictory ways. Specific comorbidities or conditions increase the risk of dying from COVID-19, such as cancer (Venkatesulu et al., 2021), cardiovascular and cerebrovascular diseases (Thakur et al., 2021), diabetes (Castro, Gurzenda, Macário, et al., 2021; Dorjee et al., 2020), hypertension (Dorjee et al., 2020), mental and behavioral disorders (Fond et al., 2021), obesity (Yang et al., 2021), pregnancy (Zambrano et al., 2020), pulmonary diseases (Dorjee et al., 2020), and renal system diseases (Dorjee et al., 2020; Thakur et al., 2021). Therefore, COVID-19 deaths may decrease the mortality from these causes, a phenomenon known as “mortality displacement” (Zeger et al., 1999). Inversely, post-Covid syndrome (known as long Covid) (Mantovani et al., 2022) has contributed to increases in mortality from cancer, cardiovascular, and respiratory diseases (Uusküla et al., 2022). Also, disruptions in the healthcare system during the pandemic, healthcare system access bias, or declining demand for healthcare from fear of contracting COVID-19 increased mortality from causes amenable to primary care (e.g., diabetes, leukemia, maternal deaths) (Bigoni et al., 2022; Dey & Davidson, 2021; Griffin, 2021; Lai et al., 2020). Finally, indirect effects on external causes are also uncertain. For example, COVID-19 lockdowns reduced traffic volume, the number of road traffic accidents, and the number of transport accident deaths. Nevertheless, some countries observed the opposite due to increased risky driving behaviors (Yasin et al., 2021).

Assessments of the impact of the COVID-19 pandemic on mortality include estimates of years of life lost (YLL), excess deaths, and changes in life expectancy at birth (𝑒_0_) and at other specific ages (e.g., 65 years). Some of these studies explore the direct (COVID-19 deaths) and indirect (non-COVID-19 deaths) mortality impacts of the COVID-19 pandemic according to different dimensions, including age, sex, race and ethnicity, socioeconomic status, regional distribution, and mortality causes (Ackley et al., 2022; Andrasfay & Goldman, 2021; Arias et al., 2021, 2022; Brant et al., 2020; Castro et al., 2022; Castro, Gurzenda, Turra, et al., 2021; Chan et al., 2021; Cronin & Evans, 2021; Guimarães et al., 2022; Iuliano et al., 2021; Jardim et al., 2022; Kelly et al., 2021; Kontopantelis et al., 2021; Lima et al., 2021; Marinho et al., 2020; Sanmarchi et al., 2021; Santos et al., 2021; Stokes et al., 2021; World Health Organization, 2022). These assessments show that along with specific mortality causes, the COVID-19 pandemic may result in decreasing indirect mortality for particular populations (e.g., economically privileged) and countries (e.g., Belgium, Canada, Costa Rica, and France). Besides socioeconomic inequalities and unequal responses to the COVID-19 pandemic, indirect or direct mortality impacts of COVID-19 may be explained by differences in health systems, COVID-19 testing, the ability to work from home, and shelter-in-place policies, and corroborate that detailed analyses by mortality causes are essential.

Here, we extend early analyses by measuring the impacts of the COVID-19 pandemic on the profile of all other causes of death in Brazil in 2020 and 2021. We have two primary goals. First, we use time series decomposition to describe changes in age-standardized mortality rates from 14 groups of causes other than COVID-19. Second, as the COVID-19 pandemic affected mortality from other causes, we measure the contribution of these 14 groups of causes of death to the annual changes in 𝑒_0_ from 2017 to 2021. Recognizing the spatial heterogeneity of the pandemic in Brazil (Castro, Kim, et al., 2021) and the distinct mortality pattern by age and for males and females, we detail the analyses by region, major age groups, and sex.

## Materials and Methods

### Population Data

We use mid-year population (July 1^st^) estimates and projections from the Brazilian Institute of Geography and Statistics (IBGE), from 2010 to 2022, for the five Brazilian regions (North, Northeast, Midwest, Southeast, and South), by five-year age groups (0-4 to 90+), and sex (male and female) (IBGE, 2018). We apply cubic splines (Forsythe et al., 1977; Zeileis & Grothendieck, 2005) to these mid-year populations to estimate 144 mid-month populations (i.e., from January 15, 2010, to December 15, 2021) by region, five-year age groups, and sex. We compute the mid-month population totals for regions and sex by tallying their respective categories (Appendices, Figure S1).

### Mortality Data

We use death registers from the Mortality Information System (SIM) of the Brazilian Ministry of Health from January 1^st^, 2010, to December 31^st^, 2021, comprising 15,847,407 records (Brasil, 2022a, 2022b). We exclude records with missing information on sex (n=7,289), age (n=38,645), or indicating COVID-19 as the cause of death but with the date of death before the year 2020 (n=6). Since the exclusion criteria overlap, the final database consists of 15,804,832 records. For each Brazilian region, we classify deaths by month, five-year age groups (0-4 to 90+), sex, and 14 groups of causes of death: diabetes mellitus; digestive system; heart and stroke; hypertension and hypertensive renal; ill-defined and unknown; influenza, pneumonia, chronic and other lower respiratory; kidney and urinary system; malignant neoplasms; mental and behavioral disorders; obesity; pregnancy, childbirth, and puerperium; septicemia; remaining causes; and COVID-19 (Appendices, Table S1).

### COVID-19 Deaths

In the first months of 2020, without specific guidelines, Medical Doctors in Brazil categorized most COVID-19 deaths using the International Classification of Diseases (ICD-10) code U04.9, severe acute respiratory syndrome (SARS). In April 2020, the World Health Organization (WHO) published guidelines for the certification and classification of COVID-19 infection as a cause of death (World Health Organization, 2020), including two new ICD-10 codes: U07.1 – COVID-19 virus identified – and U07.2 – COVID-19 virus not identified. In April 2020, the Brazilian Ministry of Health determined using ICD-10 code B34.2 for COVID-19 deaths. Prior COVID-19 deaths categorized as U04.9, U07.1, or U07.2 should be corrected to B34.2 (Brasil, 2020, 2021a, 2021b). The SIM database contains 9,278 records for 2021, with U04.9, U07.1, or U07.2 as the cause of death. Consequently, we classify all death registers with the underlying cause of death equal to B34.2, U04.9, U07.1, or U07.2 as COVID-19 deaths.

### Decomposition of Monthly Time Series of Standardized Mortality Rates

After estimating the mid-month population and obtaining the mortality data, we calculated 144-month (𝑡) (i.e., from January 2010 to December 2021) time series mortality rates (𝑚_𝑡_) by region (Brazil and the five regions), major age groups (0-4, 5-19, 20-64, and 65+), sex (male and female), and groups of causes of death. Each of the 672 distinct time series combines the six geographical dimensions, four major age groups, two categories of sex, and 14 groups of causes of death. To keep the region-age-sex-cause specific 𝑚_𝑡_ additive within each region (i.e., they add to the region’s mid-month crude death rate), we use the mid-month population for both sexes in each region and age group as denominators. We also annualize the mid-month population (i.e., adjust by a factor of 30/365). Then, we use the total population for Brazil on July 1^st^, 2021, as the standard to estimate the corresponding 144-month time series of standardized (age-adjusted) mortality rates 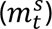.

We use the Seasonal-Trend Decomposition based on Loess (STL) method (Cleveland et al., 1990) to decompose each 144-month 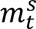 time series into three additive components: trend 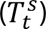, reflecting long-term changes, seasonal 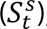, capturing intra-year fluctuations, and the remainder 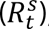, which echoes fluctuations in the data not explained by the first two components.

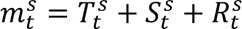

One advantage of the STL method is that it is robust to outliers (Cleveland et al., 1990), which is essential to our proposition that any mortality impacts of the COVID-19 pandemic will reflect neither on the trend nor on the seasonal components but the remainder. We specify the trend window as the total number of observations (144 months) to ensure that the trend will be as smooth as possible and reflect only long-term variations in 𝑚^#^. Another advantage of the STL method is that it allows the seasonal component to change over time (Cleveland et al., 1990), i.e., the seasonal component values for each month are not fixed. Here, we assume the season is a yearly cycle (12 months) (Rau, 2007). For COVID-19, we assume that the trend and seasonal components are equal to zero. Because both the STL method and the mortality rates are additive, within each geographical dimension, we compute the trend, seasonal and remainder totals for age groups, sex, and causes of death by adding their respective categories.

### Decomposition of Changes in Life Expectancy at Birth by Causes of Death

From the mid-year population on July 1^st^ and annual mortality data, we build annual life tables from 2017 to 2021 by region, five-year age groups (0-4 to 90+), and sex. From the same data, we also estimate annual mortality rates 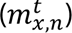 by region, five-year age groups (0-4 to 90+), sex, and groups of causes of death. To compute region-age-sex-cause specific additive decompositions (i.e., the region-age-sex-cause contributions to the changes in life expectancy at birth add to the region total change), the denominators of each region-age-sex-cause specific mortality rates are the mid-year population for both sexes in each region and age group.

We then decompose the contribution of each group of the cause of death to the annual changes in total life expectancy at birth (𝑒_0_) (i.e., 2017-2018, …, 2020-2021). We use an approach proposed by Murthy (Approach III) (Murthy, 2005), extending a method initially introduced by Pollard (Pollard, 1982). The benefit of Murthy’s approach is that it leads to minimum interaction effects between mortality changes at different ages. The contribution (main effects) of each age group 𝑥 to 𝑥 + 𝑛 to the total difference in life expectancy at birth between periods 𝑡 and 𝑡 + 1 is estimated by

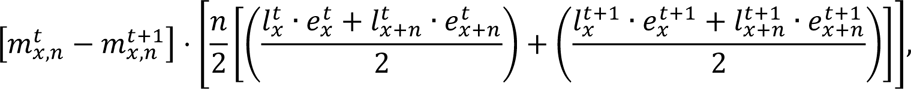

the contribution of the open age group (𝑥 +) is given by

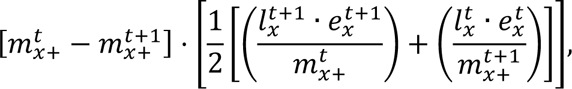

and the interaction effects for all age groups are estimated via

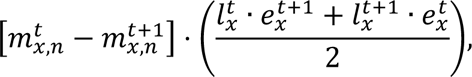

where 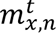 is the region-age-sex-cause specific death rate in age group 𝑥 to 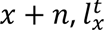 is the number of people alive at age 𝑥, and 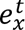 is the life expectancy at age 𝑥. Murthy (Murthy, 2005) argues that using approximate discrete equations to Pollard’s continuous method, the decomposed totals could not precisely match the observed period changes in 𝑒_0_. Yet, our estimates have minor differences constricted to around 0.5%.

## Results

We use mortality records from the Ministry of Health in Brazil to obtain a time series of deaths by cause between January 1^st^, 2010, and December 31^st^, 2021 (144 months). To comprehensively assess time trends, we estimate monthly age-standardized mortality rates for Brazil and its regions (North, Northeast, Midwest, Southeast, and South) by major age groups (0-4, 5-19, 20-64, 65+), sex, and 14 groups of underlying causes of death (Materials and Methods). The reader is cautioned that the scales of the panels included in our figures are different to uncover variations from each cause of death.

After the onset of the COVID-19 pandemic, monthly age-standardized mortality rates for all other groups of causes of death changed their temporal trajectories (Figure 1). Mortality rates for pregnancy, childbirth and puerperium, diabetes mellitus, and hypertension and hypertensive renal diseases increased following the pattern of the two major pandemic waves in Brazil in 2020 and 2021. Rates for mental and behavioral disorders reverted their trend of decline. Besides, the typical fluctuations in rates of influenza, pneumonia, and chronic and other lower respiratory diseases were not observed anymore. Also, deaths from heart and stroke decreased on average by 500 per year between 2017 and 2019, then fell by 15,000 in 2020 and increased by 15,000 in 2021 (Appendices, Tables S2 and S3). The pattern of those changes was not uniform across regions (Appendices, Figure S4).

**Figure 1.**
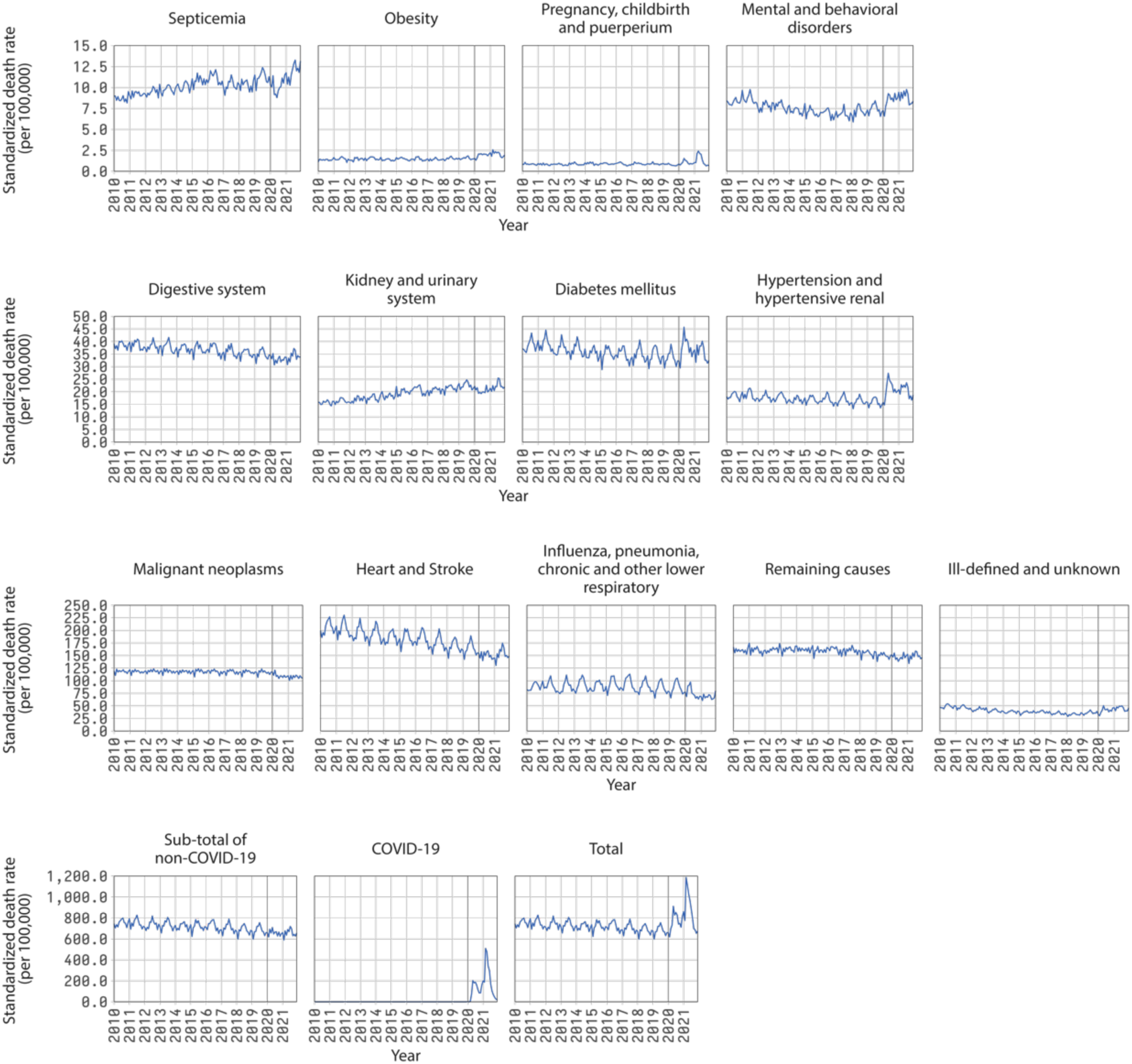
Time series of monthly standardized death rates by groups of causes of death, Brazil, 2010-2021. The reader is cautioned that the vertical scales of the panels are different to uncover variations from each cause of death.

To better evaluate the effect of the COVID-19 pandemic on other causes of death, we decompose each standardized mortality rate time series into three additive components: trend, seasonality, and the remainder (Materials and Methods). Given the long-term trend and historical seasonal fluctuations (i.e., signal) in standardized mortality rates, we assume that any impact from the pandemic (i.e., noise) will be left on the remainder component.

Figure 2 shows the remainder of the decomposition of standardized mortality rates for Brazil and both sexes (the trend and seasonal components are presented in Appendices, Figures S2 and S3, respectively). The pattern of the remainder component in 2020 and 2021 mirrors the two major pandemic waves, except for obesity and mental and behavioral disorders. However, the direction (increase or decline) and the magnitude of the change by pandemic wave varied by cause of death. Notable increases are observed for causes related to pregnancy, childbirth and puerperium, obesity, mental and behavioral disorders, diabetes mellitus, hypertension and hypertensive renal, and ill-defined and unknown causes. By contrast, groups of causes related to the digestive system, kidney and urinary system, malignant neoplasms, heart and stroke, influenza, pneumonia, chronic and other lower respiratory diseases, and remaining causes declined. Malignant neoplasms, influenza, pneumonia, chronic, and other lower respiratory diseases were uniformly affected by both COVID-19 pandemic waves.

**Figure 2.**
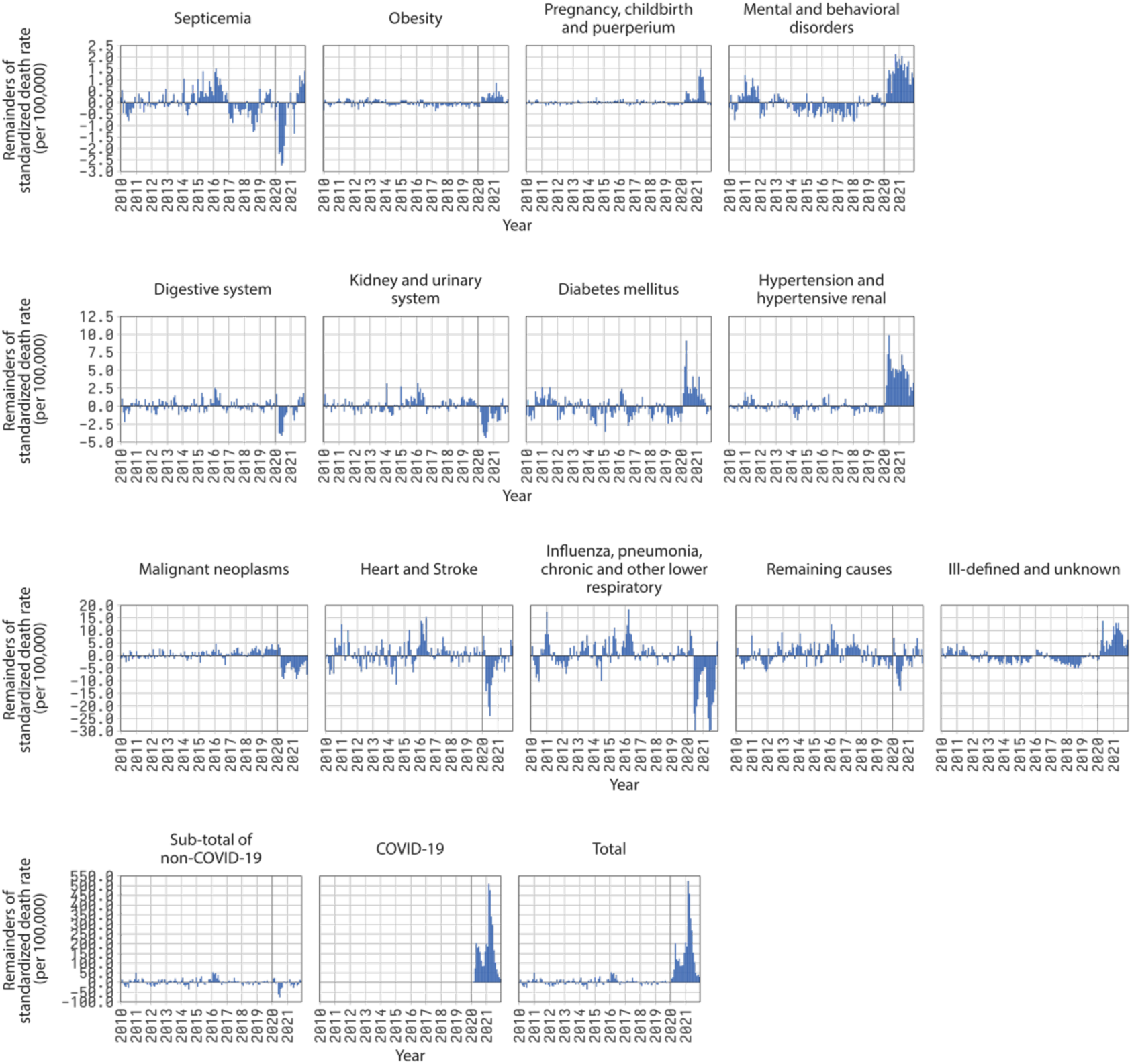
Remainder component of the decomposition of time series of monthly standardized death rates by groups of causes of death, Brazil, 2010-2021. Each graph shows the sum of the remainder obtained for each major age group. The graph for “Total” is the sum of the remainder component for each group of causes of death. The reader is cautioned that the vertical scales of the panels are different to uncover variations from each cause of death.

There are noteworthy regional differences in the levels and patterns of the pandemic impacts. Figure 3 shows the remainder of the decomposition of standardized mortality rates for each region and sex (Appendices, Figures S5 and S6 show the trend and seasonal components, respectively). Increases in the remainder for septicemia were substantial in the Midwest region in 2021, while the North region had the most significant increase in obesity. The Northeast, followed by the Southeast, had considerable increases in the remainder for mental and behavioral disorders. The most sizeable increases in pregnancy, childbirth, and puerperium were in the North, Midwest, and South regions. The remainder for diabetes mellitus, hypertension, and hypertensive renal increased substantially in the North and Northeast regions. For ill-defined and unknown causes, the North had the highest remainder increases.

**Figure 3.**
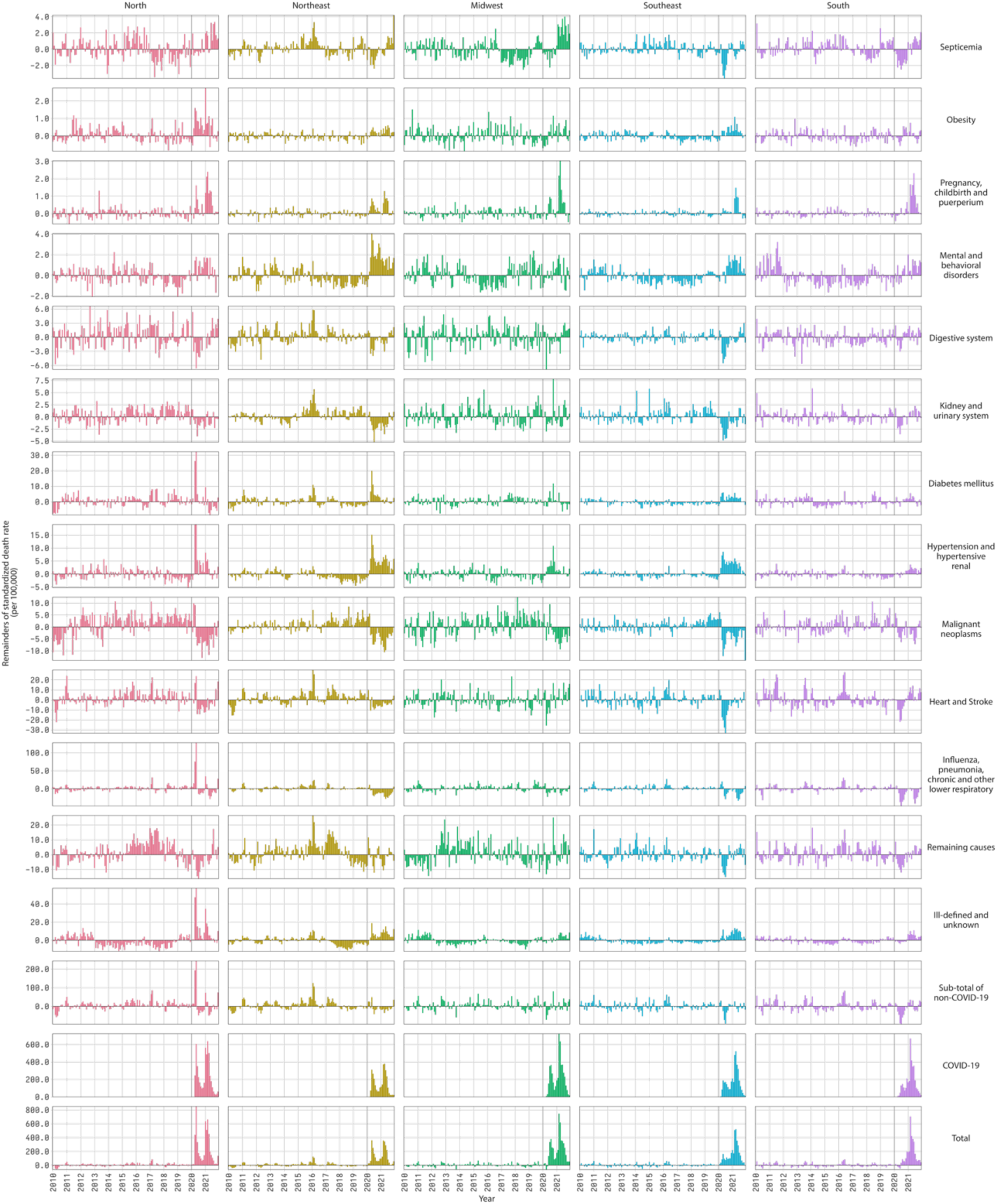
Remainder component of the decomposition of time series of monthly standardized death rates by groups of cause of death and regions, Brazil, 2020-2021. Each graph shows the sum of the remainder obtained for each major age group. The graphs for “Total” are the sum of the remainder component for each group of causes of death. The reader is cautioned that the vertical scales of the panels are different to uncover variations from each cause of death.

Considering males and females (Appendices, Figure S7), the most striking difference in the remainder component (Figure 4) is for mental and behavioral disorders, which was negligible for women but high throughout the pandemic for men. For all other causes, the levels and patterns by gender were roughly similar (Appendices, Figures S8 and S9 show the trend and seasonal components, respectively).

**Figure 4.**
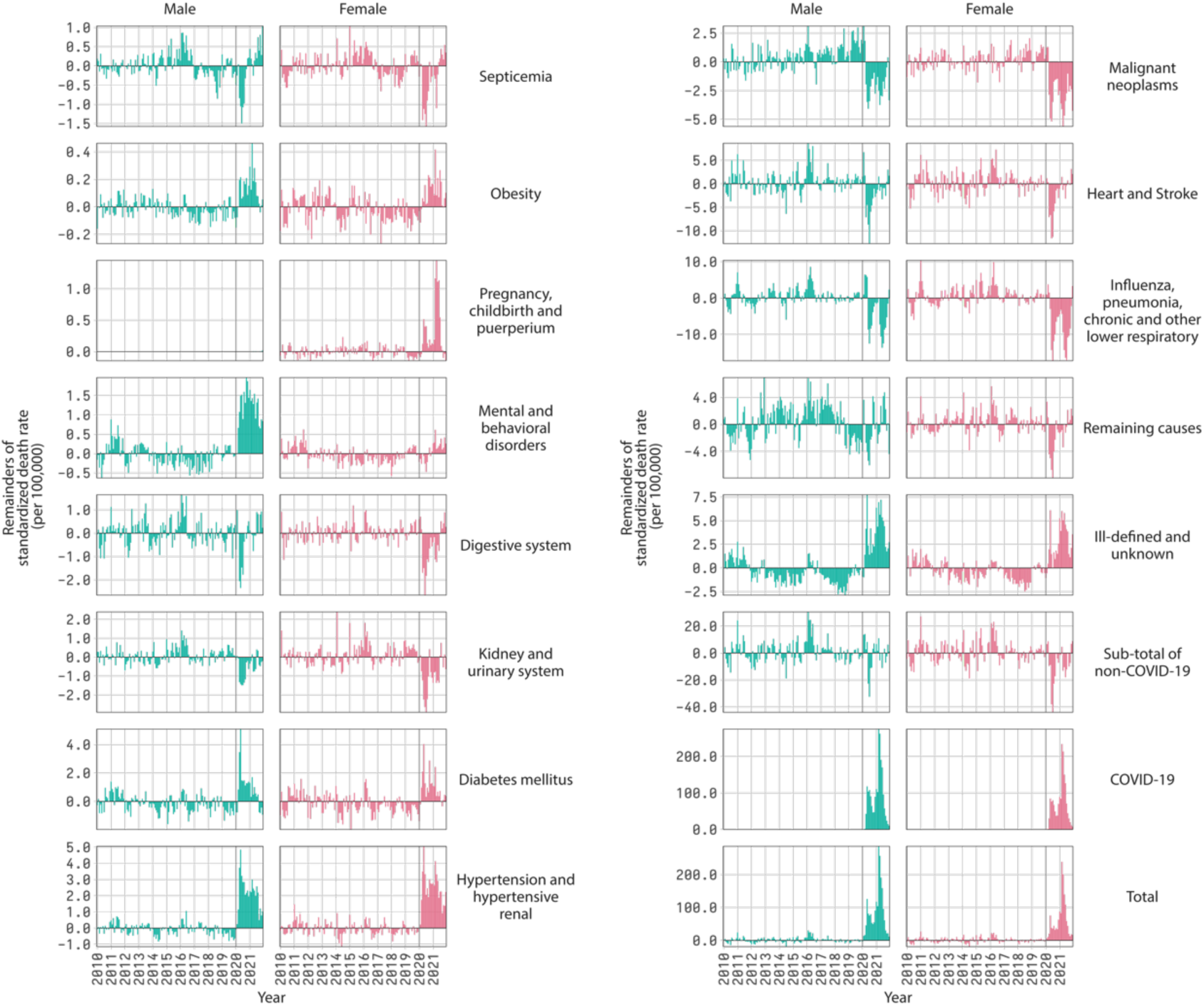
Remainder component of the decomposition of time series of monthly standardized death rates by groups of causes of death and sex, Brazil, 2020-2021. Each graph shows the sum of the remainder obtained for each major age group. The graphs for “Total” are the sum of the remainder component for each group of causes of death. The reader is cautioned that the vertical scales of the panels are different to uncover variations from each cause of death.

Concerning the major age groups 20-64 and 65+ (Appendices, Figures S10 to S13), the marked difference in the remainder component (Appendices, Figure S13) is for mental and behavioral disorders, which was three times higher for 20-64. For septicemia, digestive system, and remaining causes, the remainder component presented an inverted direction of change, positive for 20-64 and negative for 65+. For all other causes, except for obesity, the age group 65+ showed a more significant change in levels than 20-64.

Given the different ways the COVID-19 pandemic affected causes of death, we decompose the contributions of COVID-19 deaths (direct effect) and the other groups of causes (indirect effects) to the annual change in the observed life expectancy at birth (𝑒_0_) in Brazil from 2017 to 2021 (Materials and Methods). During 2020 and 2021, the COVID-19 pandemic reduced 𝑒_0_ in Brazil by 3.23 years, 1.44 in 2020, and 1.79 in 2021 (Appendices, Table S4). Direct effects reduced 𝑒_0_ by 1.89 years in 2020 and 1.77 in 2021. Indirect effects increased 𝑒_0_ by 0.44 in 2020 and had virtually no impact on 𝑒_0_ in 2021 (0.01 years).

Regarding specific causes of death, our findings are consistent with the change in the pattern of the remainder component (Figure 5). Causes of death with significant increases in the remainder (i.e., obesity; pregnancy, childbirth, and puerperium; mental and behavioral disorders; diabetes mellitus, hypertension, and hypertensive renal disease; and ill-defined and unknown) contributed to reducing 𝑒_0_ by 0.27 years in 2020 and 0.05 years in 2021. Causes of death with decreases in the remainder (i.e., digestive system; kidney and urinary system; malignant neoplasms; heart and stroke; influenza, pneumonia, chronic and other lower respiratory diseases; and remaining causes) increased 𝑒_0_ by 0.69 years in 2020 and 0.06 years in 2021. Septicemia, which had mixed impacts from the pandemic, increased 𝑒_0_ by 0.02 in 2020 and decreased 𝑒_0_ by 0.03 in 2021.

**Figure 5.**
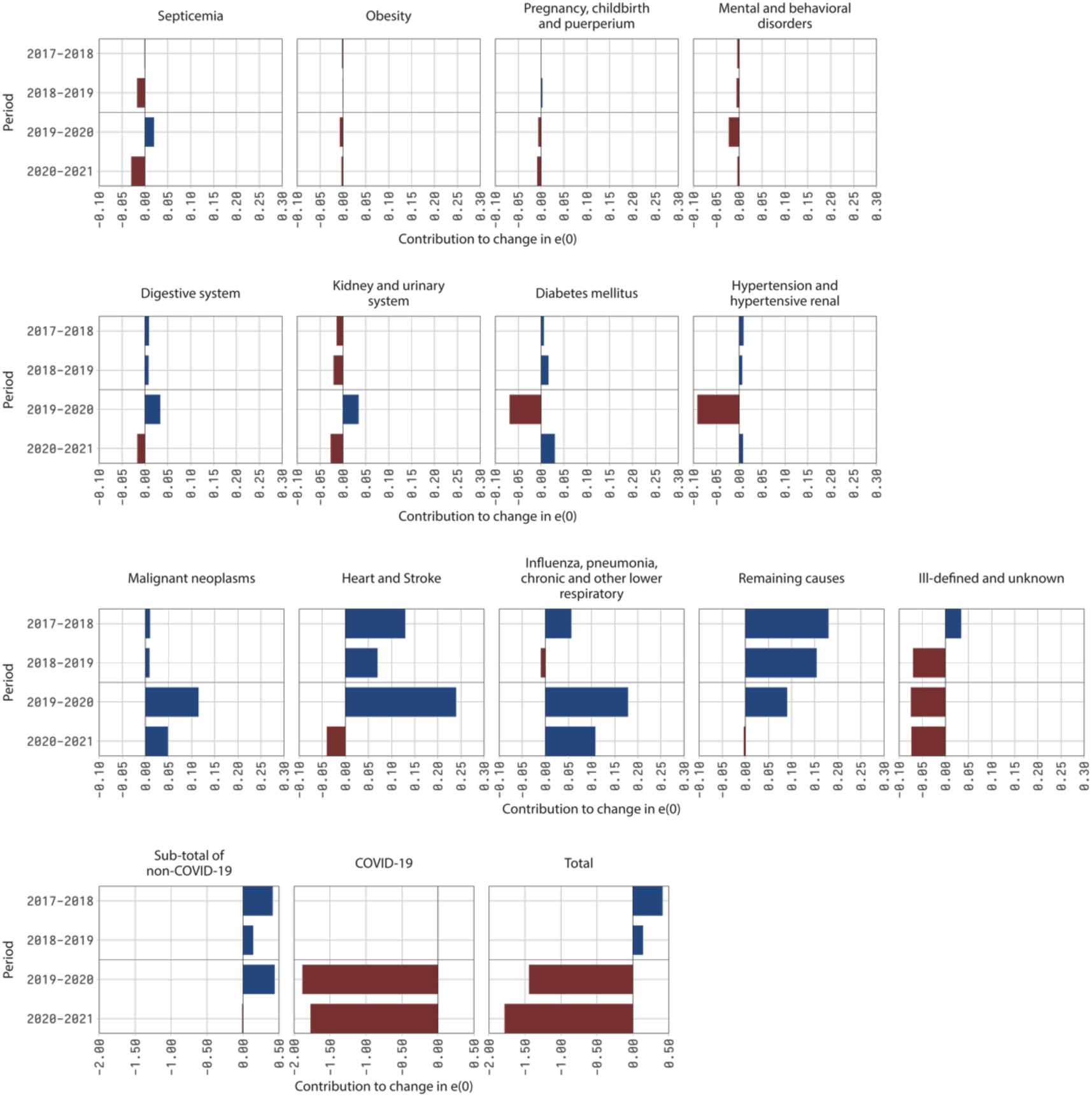
Contribution of groups of causes of death to the change in life expectancy at birth (𝑒_0_), Brazil, 2017-2021. The graph for “Total” is the sum of the contribution of each cause of death. The reader is cautioned that the horizontal scales of the panels are different to uncover variations from each cause of death.

Concerning regions and sex (Figure 6 and Appendices, Table S5), the largest declines in 𝑒_0_ in 2020 and 2021, respectively, occurred in the North (2.79 years) and South (3.32 years) regions. In both cases, the impact was higher for males. Combining 2020 and 2021, the highest reduction in 𝑒_0_ from the COVID-19 pandemic was in the Midwest (4.30 years, 1.85 in 2020, 2.46 in 2021) and the lowest in the Northeast (2.42 years, 1.87 in 2020, 0.54 in 2021). Direct effects reduced 𝑒_0_ the most in the Midwest (4.62 years, 2.21 in 2020, 2.41 in 2021) and the least in the Northeast (2.60 years, 1.84 in 2020, 0.77 in 2021). On the contrary, in 2020 and 2021 combined, the indirect effects of the COVID-19 pandemic increased 𝑒_0_ in all regions. Indirect effects increased 𝑒_0_ in 2020 in the Midwest, Southeast, and South, and 2021 in the North and Northeast. Indirect effects increased 𝑒_0_ the most in the South in 2020 (0.94 years) but contributed to the largest reduction in 𝑒_0_ in the same region in 2021 (0.50 years).

**Figure 6.**
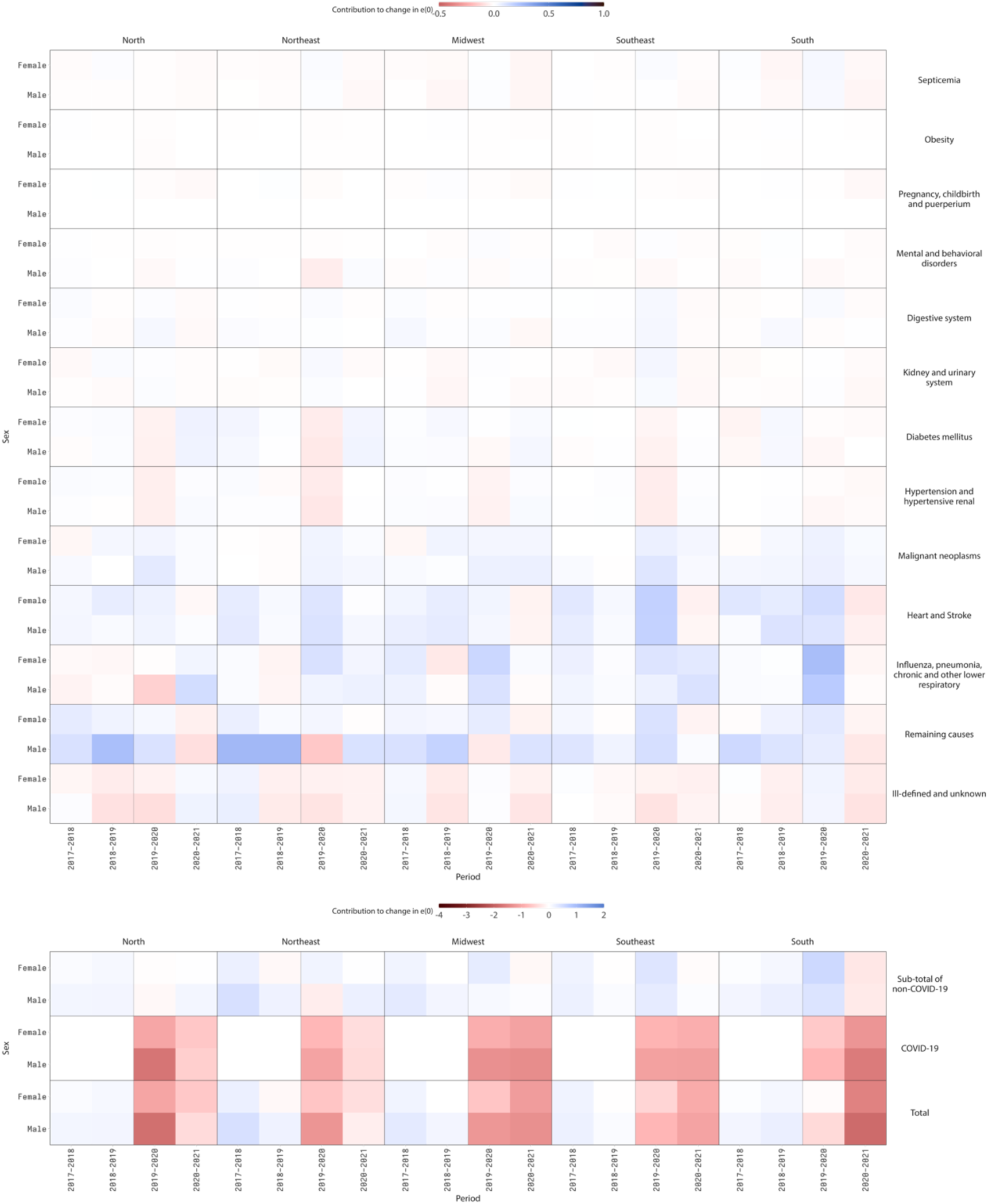
Contribution of groups of causes of death to the change in life expectancy at birth (𝑒_0_) by region and sex, Brazil. The values for “Total” are the sum of the contribution of each cause of death. The reader is cautioned that the color scales of the panels are different to uncover variations from each cause of death.

Considering regions and specific causes of death (Appendices, Table S5), the most relevant increases from indirect effects in 𝑒_0_ came from influenza, pneumonia, chronic and other lower respiratory diseases in all regions except in the North in 2020 and the South in 2021, malignant neoplasms in all regions in 2020 and 2021, heart and stroke in all regions in 2020, and remaining causes in the North, Southeast, and South in 2020. By contrast, sharp decreases from indirect effects in 𝑒_0_ came from hypertension and hypertensive renal and diabetes mellitus in all regions in 2020, ill-defined and unknown in all regions except in the Midwest and the South in 2020 and the North in 2021, influenza, pneumonia, chronic and other lower respiratory diseases in the North in 2020, remaining causes in the Northeast in 2020 and the North in 2021, and heart and stroke in the South in 2021.

There are also different contributions to changes in 𝑒_0_ by major age groups (Figure 7 and Appendices, Table S6). Except for a few regions, changes in mortality caused by influenza, pneumonia, chronic and other lower respiratory diseases (ages 65+), remaining causes (ages 65+ in 2020), heart and stroke (ages 20-64 and 65+ in 2020), and malignant neoplasms (ages 20-64 and 65+) increased life expectancy at birth. Conversely, relevant decreases from indirect effects in 𝑒_0_ came from remaining causes (ages 20-64), ill-defined and unknown causes of death (ages 20-64 and 65+), hypertension and hypertensive renal diseases, and diabetes mellitus (ages 20-64 and 65+ in 2020).

**Figure 7.**
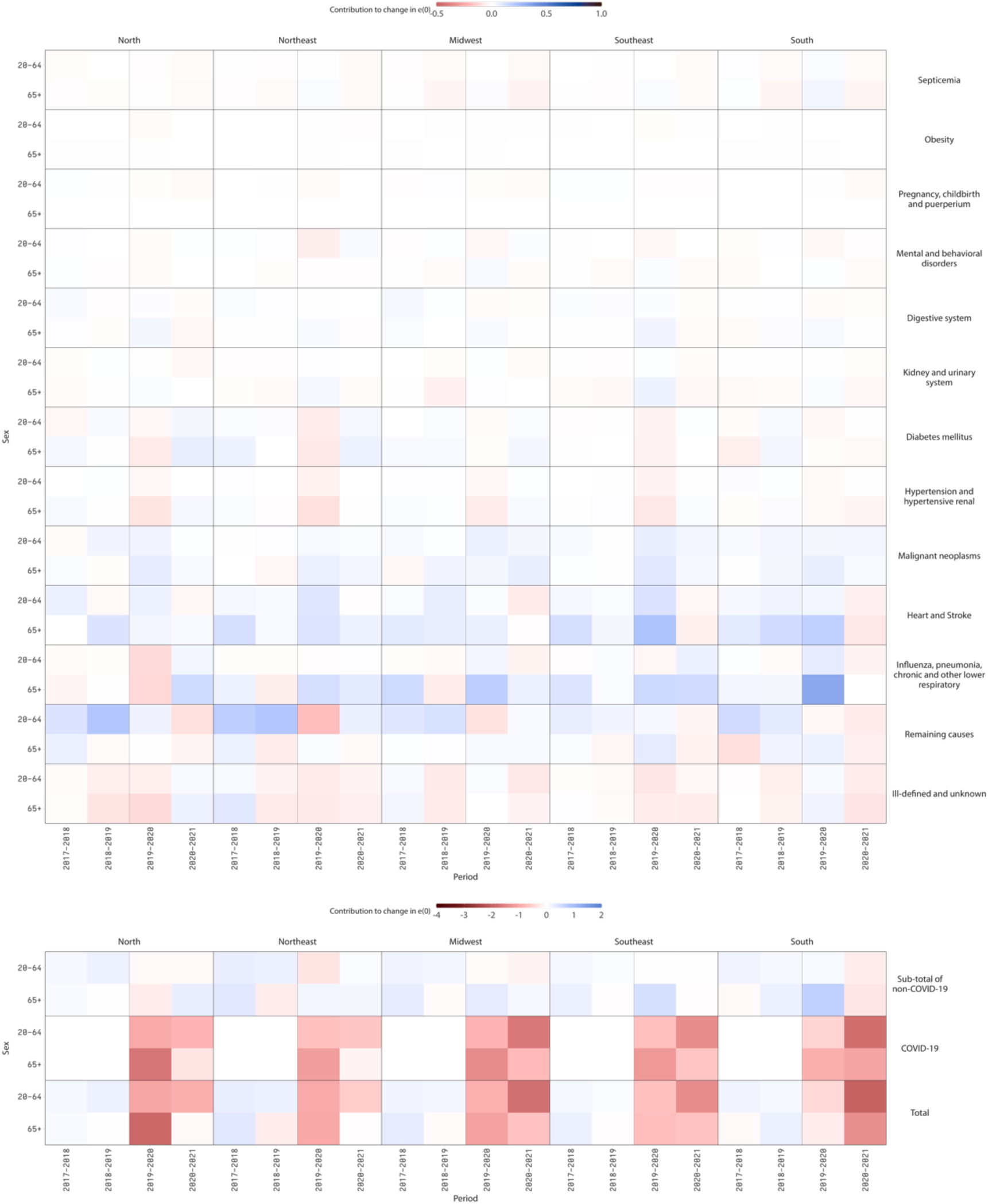
Contribution of groups of causes of death to the change in life expectancy at birth (𝑒_0_) by region and major age groups, Brazil. The values for “Total” are the sum of the contribution of each cause of death. The reader is cautioned that the color scales of the panels are different to uncover variations from each cause of death.

## Discussion

This study aimed to examine the impact of the COVID-19 pandemic on other causes of death in Brazil in 2020 and 2021. Our results show that consequences were distinct in direction and magnitude when detailed by causes of death, age groups, sex, and geographical regions. Broadly, the remainder component of the time series of mortality rates from groups of causes of death mirrors the pattern of the two major pandemic waves. However, the impact of the COVID-19 pandemic on other causes of death was not limited to increases but decreases as well. As a result, the direct effects of the COVID-19 pandemic in Brazil reduced 𝑒_0_ by 1.89 years between 2019 and 2020 and 1.77 between 2020 and 2021. Indirect effects increased 𝑒_0_ by 0.44 between 2019 and 2020 and had virtually no impact on 𝑒_0_ between 2020 and 2021.

The main strength of this study is the comprehensive analysis of mortality by underlying causes of death, sex, and region, using Brazil’s most complete dataset of mortality records. The limitations are mostly related to the classification of the cause of death. It is possible that the misclassification of COVID-19 deaths (Gill & DeJoseph, 2020; Moghadas & Galvani, 2021) resulted in an overestimation of our results for specific causes of death. Additionally, misclassifying other causes and underreporting of deaths could have happened differently by sex and region. However, the Brazilian Ministry of Health’s April 2020 guidelines for proper ICD-10 categorization of COVID-19 deaths (Brasil, 2020, 2021a, 2021b) (Materials and Methods) have minimized this problem. Also, underreporting of deaths in Brazil is low (around 1.5%) (IBGE, 2022), and ill-defined and unknown deaths have remained unchanged since 2019 (about 5%).

On the one hand, some of our findings support previous studies which show that the COVID-19 pandemic may increase the mortality of other causes of death (Arias et al., 2021, 2022; Brant et al., 2020; Guimarães et al., 2022; Jardim et al., 2022; Kontopantelis et al., 2021; Santos et al., 2021; Stokes et al., 2021). This is consistent with the idea that disruptions in the healthcare system caused by the COVID-19 pandemic (Bigoni et al., 2022; Dey & Davidson, 2021; Griffin, 2021; Lai et al., 2020) increased the mortality from causes amenable to primary care (Nolte & McKee, 2004). On the other hand, our results also corroborate earlier studies that reveal that the COVID-19 pandemic may also decrease the mortality of other causes of death, such as heart and stroke (Brant et al., 2020; Jardim et al., 2022; Santos et al., 2021), influenza, pneumonia, chronic and other lower respiratory diseases (Arias et al., 2021, 2022; Guimarães et al., 2022; Kontopantelis et al., 2021; Santos et al., 2021), malignant neoplasms (Arias et al., 2021, 2022; Jardim et al., 2022), as well the digestive system, kidney, and urinary system. This is consistent with competing risks (Chiang, 1991; Yashin et al., 1986) and corroborates earlier studies that associated specific morbidities or conditions with increased risk of dying from COVID-19 (Castro, Gurzenda, Macário, et al., 2021; Dorjee et al., 2020; Fond et al., 2021; Thakur et al., 2021; Venkatesulu et al., 2021; Yang et al., 2021; Zambrano et al., 2020). Therefore, our analysis suggests that mortality replacement did happen in Brazil during the COVID-19 pandemic.

We cannot rule out the possibility that the misclassification of causes of death could explain some of the differences. The North or Northeast regions had the most significant increases or decreases of the time series remainders. These regions are marked by striking inequalities and have some of the worst socioeconomic, health outcomes, and health infrastructure and access indicators (IBGE, 2021; Rocha et al., 2021). Restricted availability of resources, physicians, and intensive care unit beds are major limiting factors (Rocha et al., 2021). They also contributed to the speed of the spread of COVID-19 and adverse mortality outcomes (Castro, Kim, et al., 2021).

Regarding 𝑒_0_, our results show the largest declines for the North and Northeast regions in 2020, while in 2021, the Midwest and South regions had the leading reductions, corroborating earlier findings (Castro, Gurzenda, Turra, et al., 2021). These results reflect the regional spread of COVID-19, with a widespread late transmission in the South and Midwest (Castro, Kim, et al., 2021). Most of the decline resulted from the direct effects of COVID-19. Therefore, it is expected that as the mortality burden of the pandemic subsides, 𝑒_0_ will progressively return to its pre-pandemic temporal trajectory.

The United Nations Population Estimates and Projections 2022 assumes Brazil will return to pre-pandemic levels in 2023 (United Nations, 2022). Here, two issues are important to consider regarding the extent to which the future pace of gains in 𝑒_0_ will be like pre-pandemic levels and with the same regional gradient. First, several studies show that patients who recovered from COVID-19 have a higher mortality risk than those who have not contracted the disease (Al-Aly et al., 2022; Uusküla et al., 2022; Xie et al., 2022). There is no systematic and nationwide information on long COVID-19 in Brazil. As of July 2022, 33.8 million people have recovered from COVID-19 (likely an underestimated number due to limited testing in the early months of the pandemic and the lack of a reporting system for self-testing), and therefore a non-negligible number could potentially suffer premature mortality. Second, extreme poverty in Brazil has increased from 4.67% to 5.74% between 2019 and 2021 (Figueiredo, 2022). It is estimated that in 2014-2016 1.9% of the total population (3.9 million) faced severe food insecurity; in 2019-2021, this number increased to 7.3% (15.4 million) (FAO et al., 2022). This scenario could also affect health outcomes and result in higher morbidity and mortality among children and adults.

Considering the impacts of the COVID-19 pandemic on mortality patterns in Brazil and the current scenario of increased poverty, the timely implementation and the characteristics of government programs aimed to promote equity and guarantee a minimum income for families’ subsistence will be critical to putting the country on a path to return to pre-pandemic mortality levels. Those programs are essential to mitigate the consequences of the pandemic.

## Data Availability

We use death registers from the Mortality Information System (SIM) of the Brazilian Ministry of Health and population data from the Brazilian Institute for Geography and Statistics (IBGE), both publicly available.

## Appendices

**Table S1.** Groups of underlying causes of death by their ICD-10 codes (World Health Organization, 2020)

| Group of underlying cause of death | ICD-10 code |
| --- | --- |
| Septicemia | ○ Septicemia (A40-A41) (Heron, 2021) |
| Obesity | ○ Obesity and other hyperalimentation (E65-E68) |
| Pregnancy, childbirth, and puerperium | ○ Pregnancy, childbirth, and puerperium (O00-O99) (Heron, 2021) |
| Mental and behavioral disorders | ○ Mental and behavioral disorders (F00-F99) |
| Digestive system | ○ Diseases of the digestive system (K00-K93)<br>○ Symptoms and signs involving the digestive system and abdomen (R10-R19) |
| Kidney and urinary system | ○ Nephritis, nephrotic syndrome, and nephrosis (N00-N07, N17-N19, N25-N27) (Heron, 2021)<br>○ Infections of the kidney (N10-N12, N13.6, N15.1) (Heron, 2021)<br>○ Other diseases of the urinary system (N30, N31, N32, N34, N35, N36, N39)<br>○ Symptoms and signs involving the urinary system (R30-R39) |
| Diabetes mellitus | ○ Diabetes mellitus (E10-E14) (Heron, 2021) |
| Hypertension and hypertensive renal | ○ Essential hypertension and hypertensive renal disease (I10, I12, I15) (Heron, 2021) |
| Malignant neoplasms | ○ Malignant neoplasms (C00-C97) (Heron, 2021) |
| Heart and Stroke | ○ Diseases of the heart (I00-I09, I11, I13, I20-I51) (Heron, 2021)<br>○ Cerebrovascular diseases (I60-I69) (Heron, 2021)<br>○ Atherosclerosis (I70) (Heron, 2021)<br>○ Aortic aneurysm and dissection (I71) (Heron, 2021) |
| Influenza, pneumonia, chronic and other lower respiratory | ○ Influenza and pneumonia (J09-J18) (Heron, 2021)<br>○ Chronic lower respiratory diseases (J40-J47) (Heron, 2021)<br>○ Acute bronchitis and bronchiolitis (J20-J21) (Heron, 2021)<br>○ Other acute lower respiratory infections (J22) (Heron, 2021)<br>○ Lung diseases due to external agents (J60-J70)<br>○ Other respiratory diseases principally affecting the interstitium (J80-J84)<br>○ Suppurative and necrotic conditions of the lower respiratory tract (J85-J86)<br>○ Other diseases of pleura (J90-J94)<br>○ Other diseases of the respiratory system (J95-J99)<br>○ Asphyxia (R090)<br>○ Respiratory arrest (R092) |
| Remaining causes | ○ All remaining ICD-10 codes |
| Ill-defined and unknown | ○ Ill-defined and unknown causes of mortality (R95-R99)<br>○ General symptoms and signs (R50-R69) |
| COVID-19 | ○ Brazil's Minister of Health codes for COVID-19: (B342, U071, U072) (Brasil, 2020, 2021a, 2021b)<br>○ Severe acute respiratory syndrome [SARS] (U04) |

**Figure S1.**
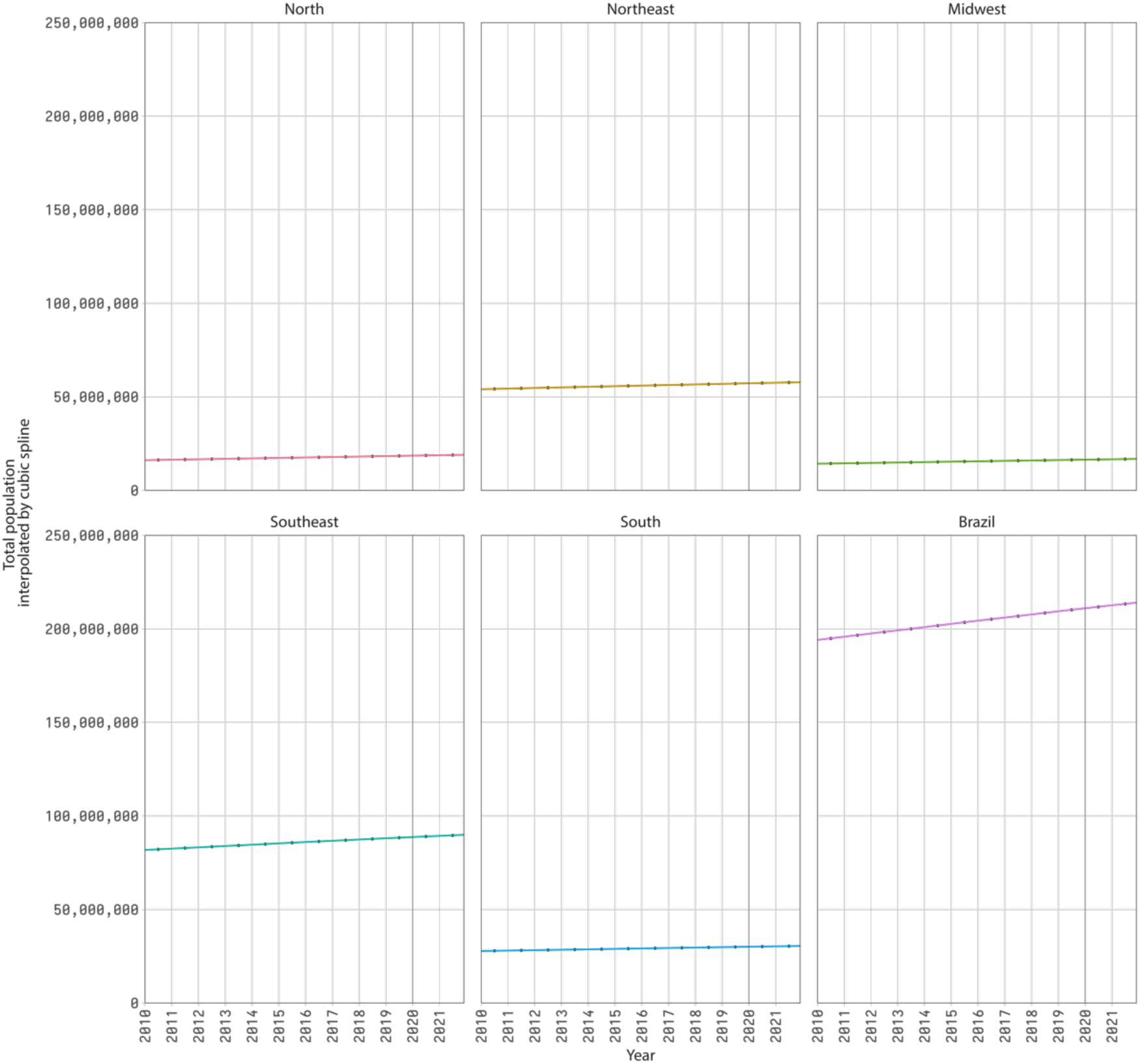
Total mid-month population (lines) from interpolating mid-year estimates and projections (dots) using cubic splines, January 15th, 2010-December 15th, 2021. The graph for “Brazil” is the sum of the population for each region.

**Figure S2.**
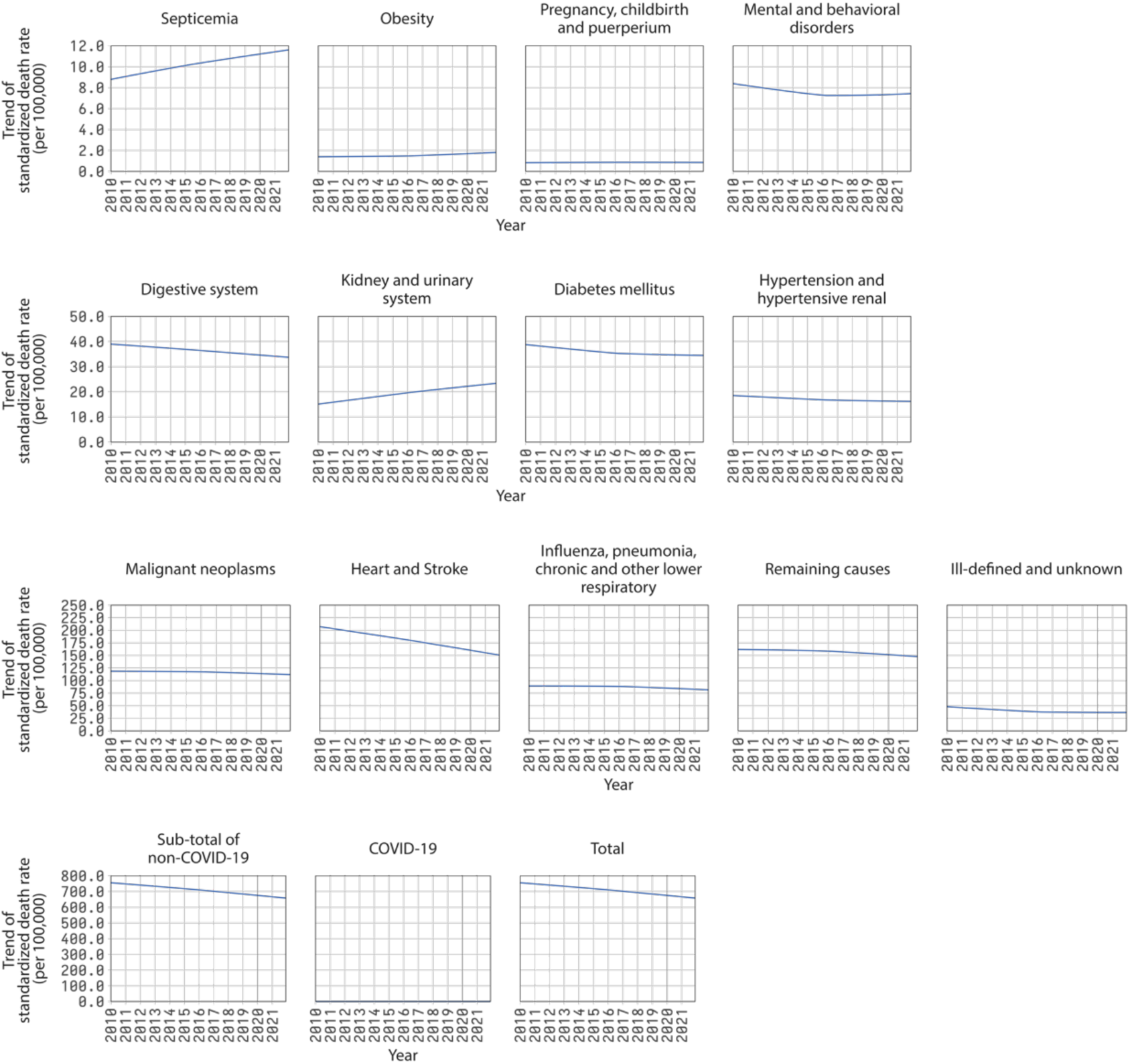
Trend component of the decomposition of time series of monthly standardized death rates by groups of causes of death, Brazil, 2010-2021. Each graph shows the sum of the trend obtained for each major age group. The graph for “Total” is the sum of the trend component for each group of causes of death. The reader is cautioned that the vertical scales of the panels are different to uncover variations from each cause of death.

**Figure S3.**
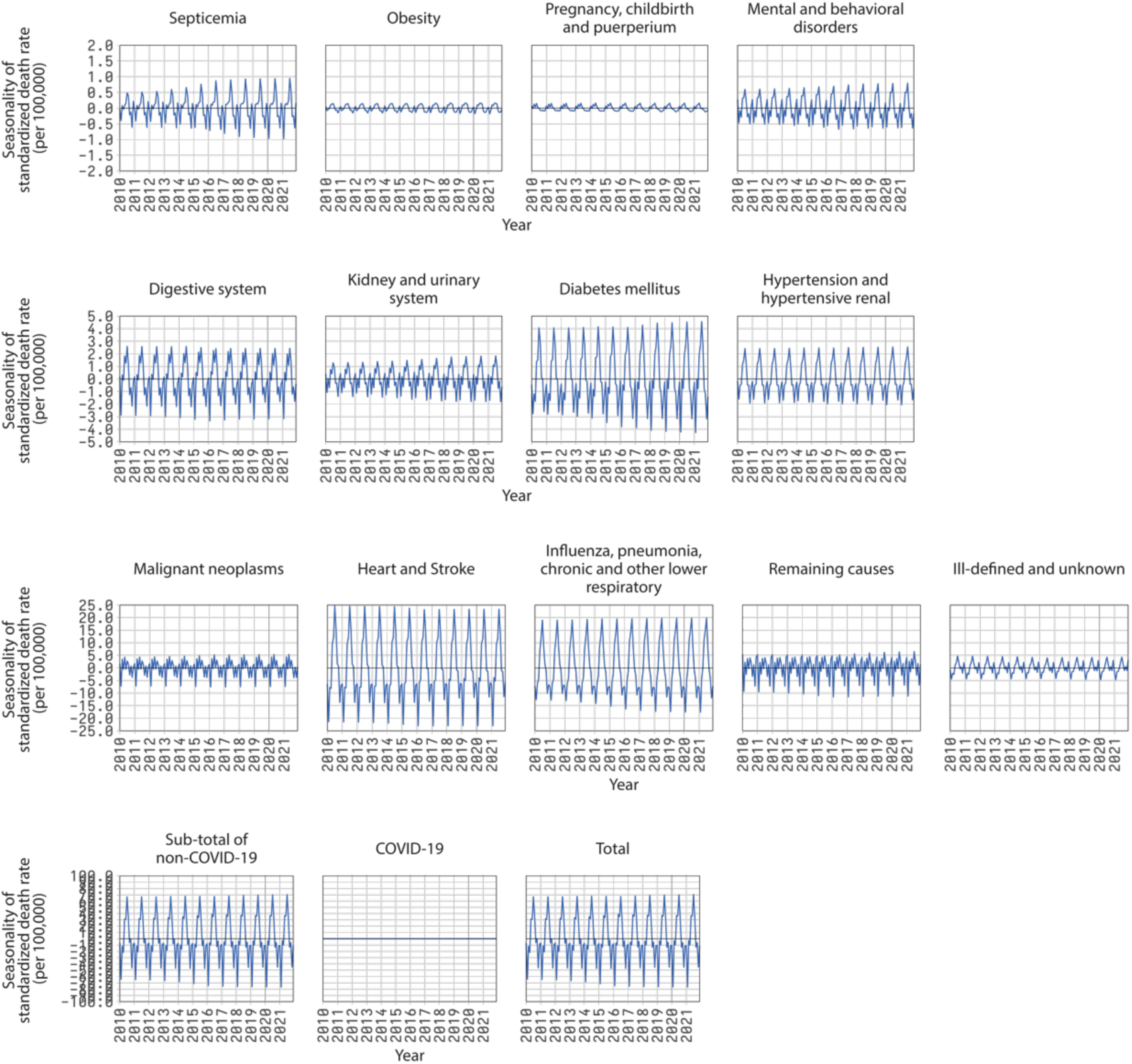
Seasonality component of the decomposition of time series of monthly standardized death rates by groups of causes of death, Brazil, 2010-2021. Each graph shows the sum of the seasonality obtained for each major age group. The graph for “Total” is the sum of the seasonality component for each group of causes of death. The reader is cautioned that the vertical scales of the panels are different to uncover variations from each cause of death.

**Figure S4.**
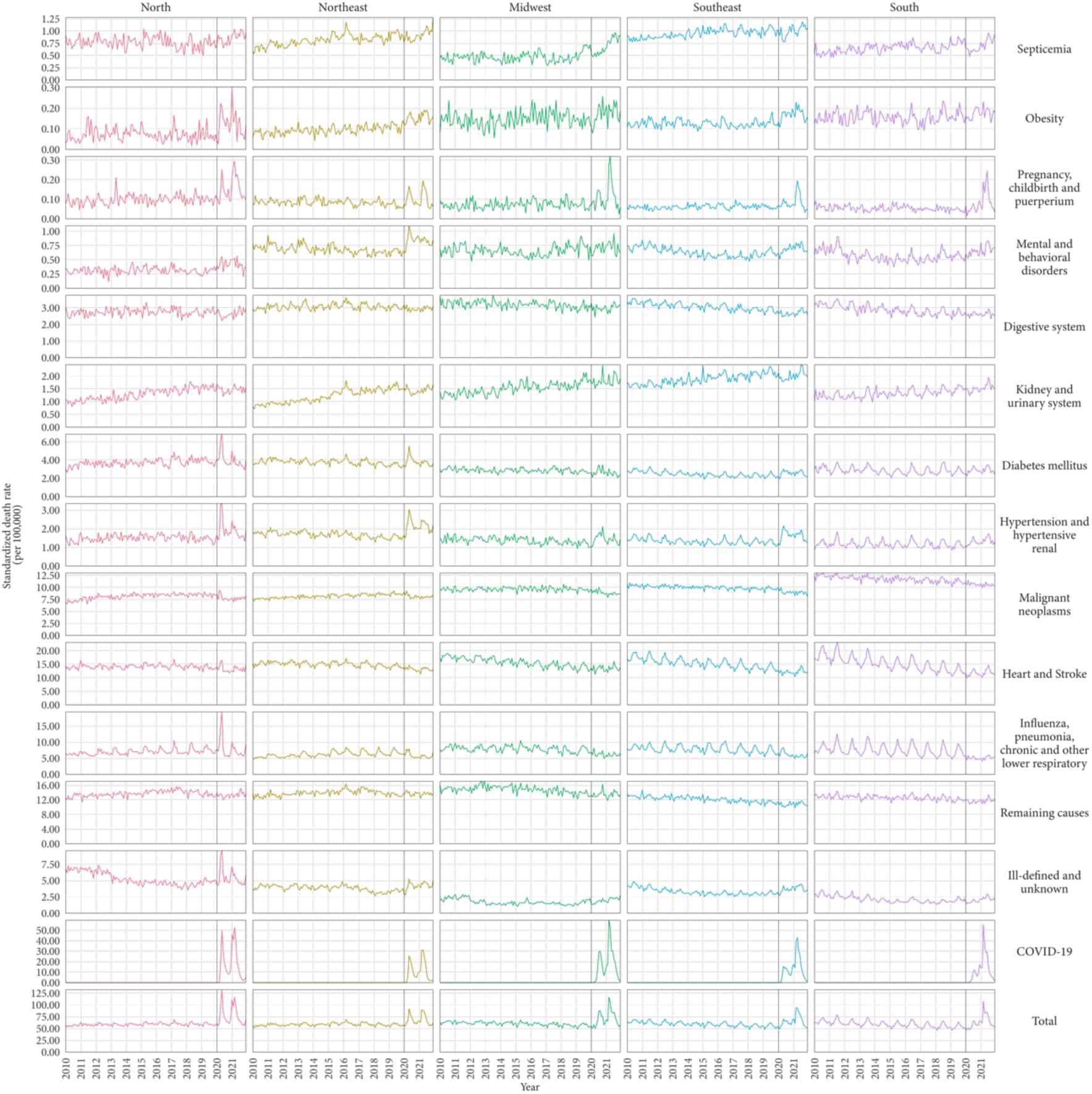
Time series of monthly standardized death rates by groups of cause of death and regions, Brazil, 2010-2021. The reader is cautioned that the vertical scales of the panels are different to uncover variations from each cause of death.

**Figure S5.**
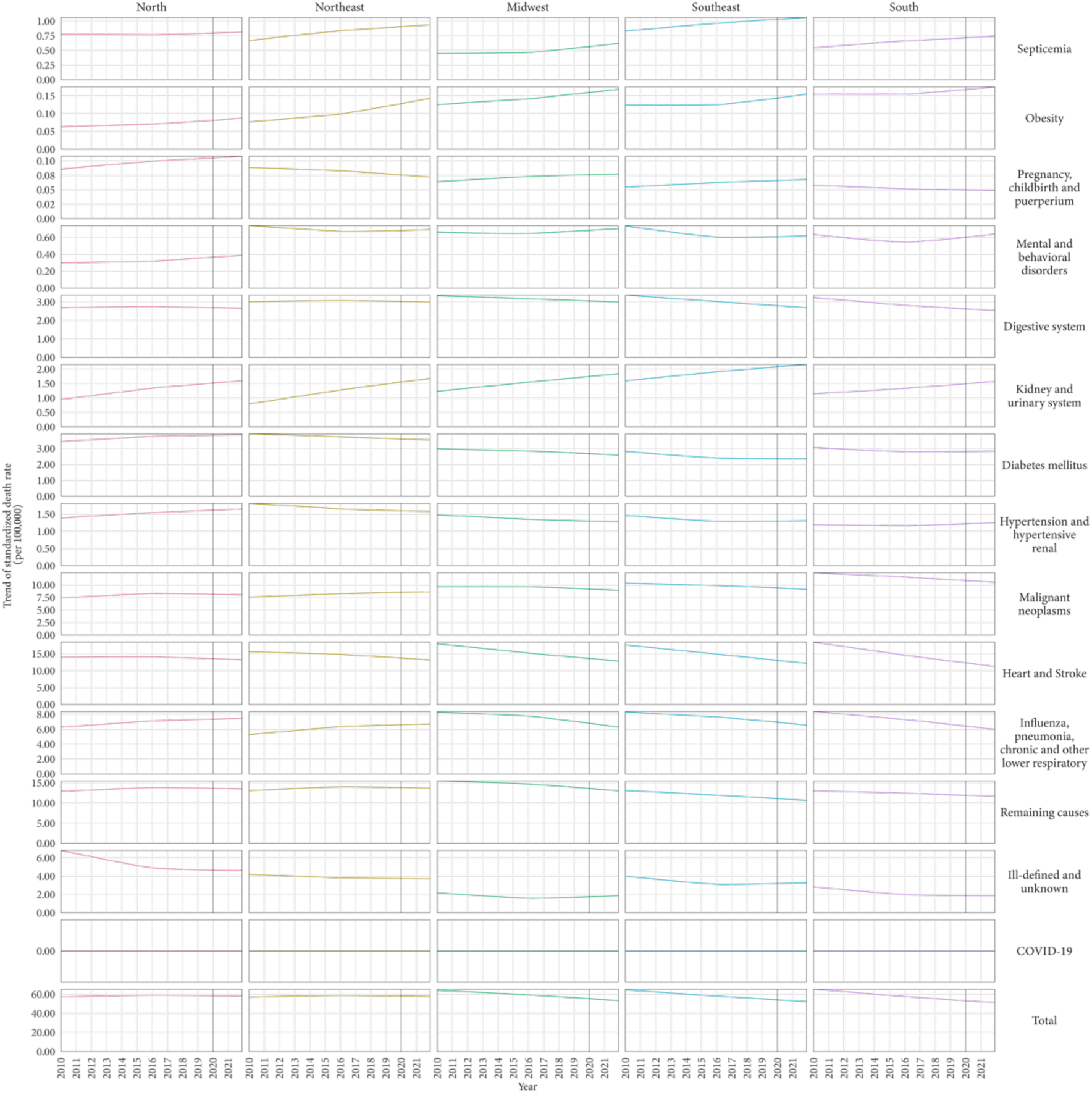
Trend component of the decomposition of time series of monthly standardized death rates by groups of cause of death and regions, Brazil, 2020-2021. Each graph shows the sum of the trend obtained for each major age group. The graphs for “Total” are the sum of the trend component for each group of causes of death. The reader is cautioned that the vertical scales of the panels are different to uncover variations from each cause of death.

**Figure S6.**
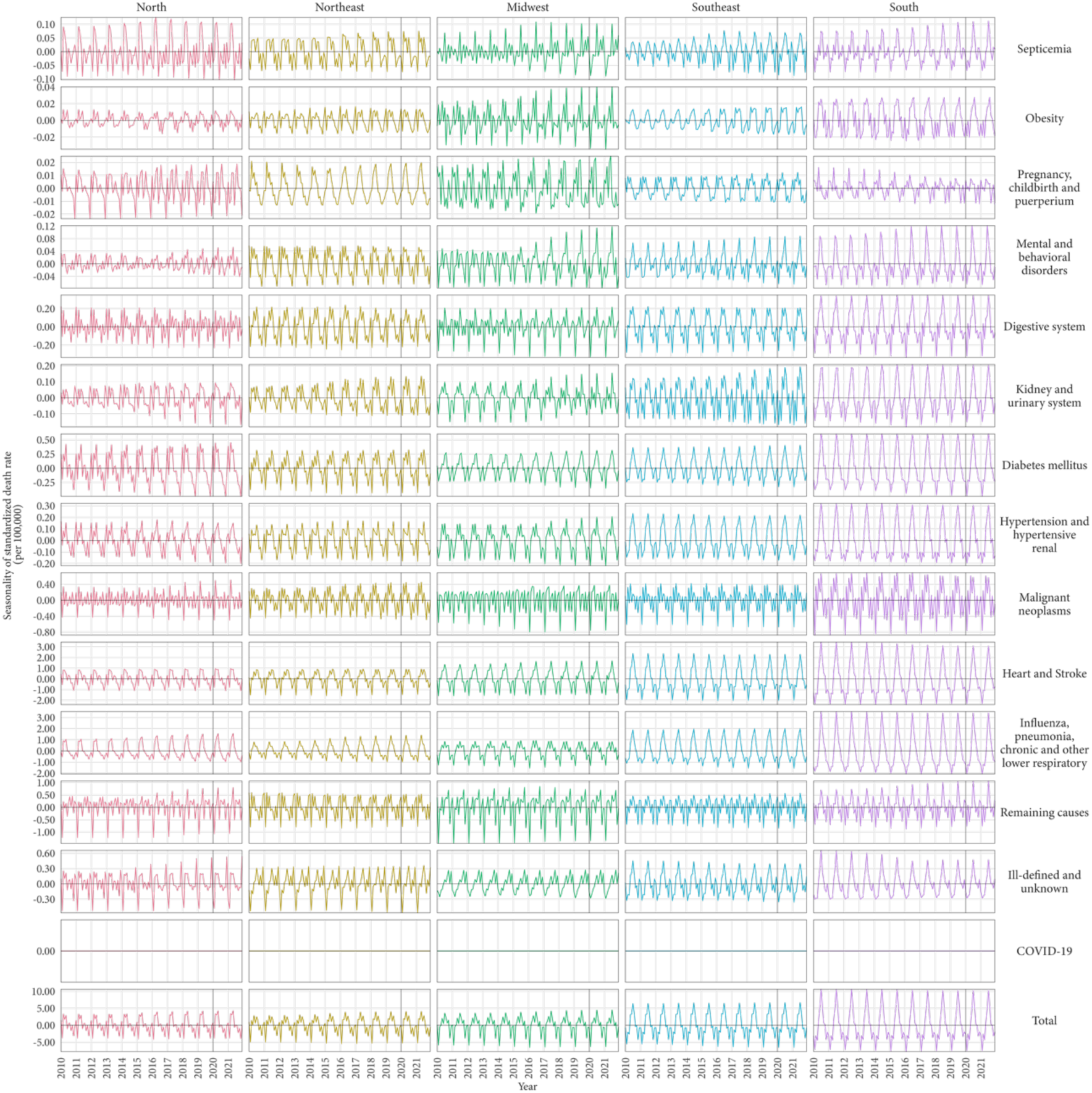
Seasonality component of the decomposition of time series of monthly standardized death rates by groups of cause of death and regions, Brazil, 2020-2021. Each graph shows the sum of the seasonality obtained for each major age group. The graphs for “Total” are the sum of the seasonality component for each group of causes of death. The reader is cautioned that the vertical scales of the panels are different to uncover variations from each cause of death.

**Figure S7.**
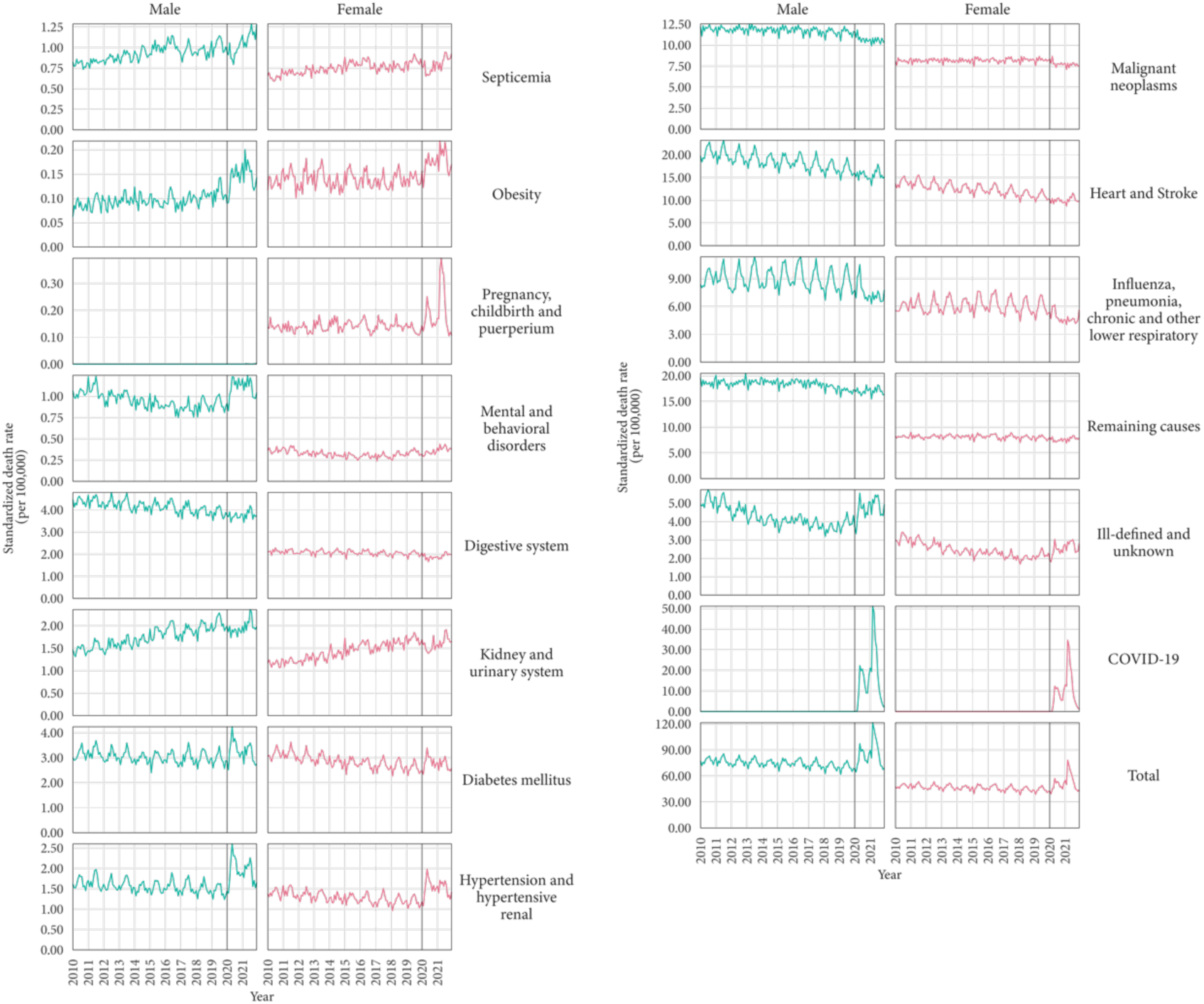
Time series of monthly standardized death rates in Brazil by groups of causes of death and sex, Brazil, 2010-2011. The reader is cautioned that the vertical scales of the panels are different to uncover variations from each cause of death.

**Figure S8.**
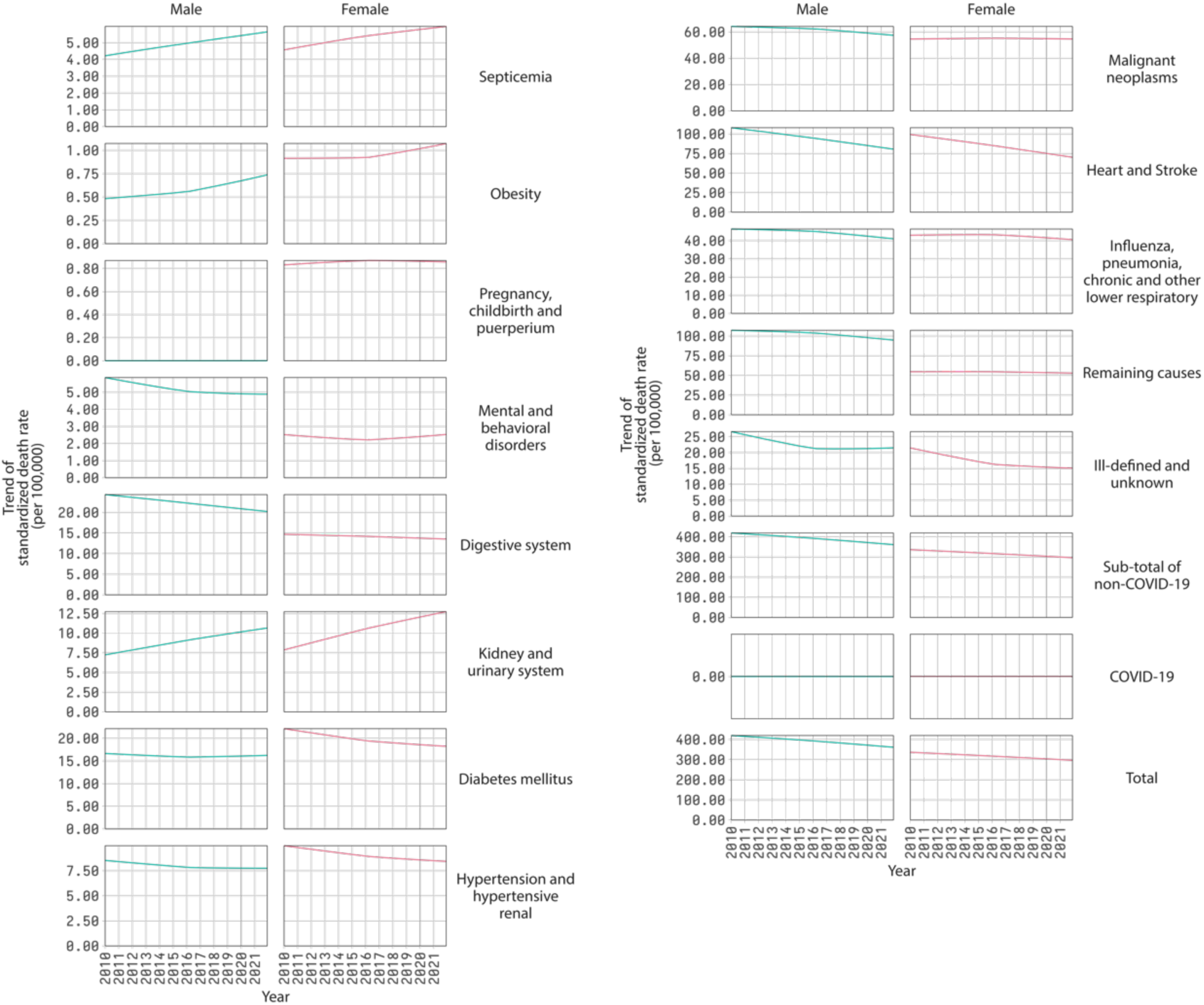
Trend component of the decomposition of time series of monthly standardized death rates by groups of cause of death and sex, Brazil, 2020-2021. Each graph shows the sum of the trend obtained for each major age group. The graphs for “Total” are the sum of the trend component for each group of causes of death. The reader is cautioned that the vertical scales of the panels are different to uncover variations from each cause of death.

**Figure S9.**
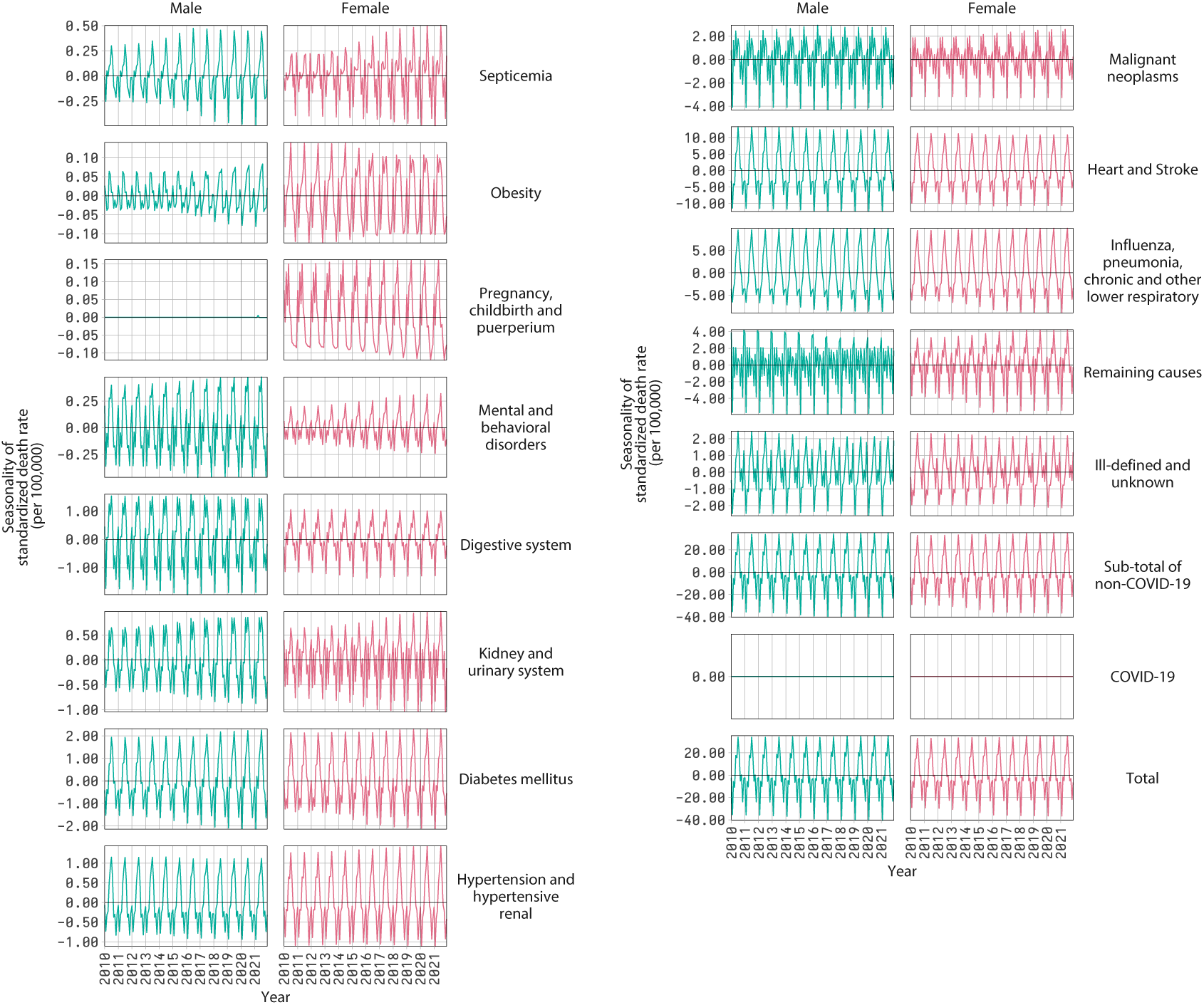
Seasonality component of the decomposition of time series of monthly standardized death rates by groups of cause of death and sex, Brazil, 2020-2021. Each graph shows the sum of the seasonality obtained for each major age group. The graphs for “Total” are the sum of the seasonality component for each group of causes of death. The reader is cautioned that the vertical scales of the panels are different to uncover variations from each cause of death.

**Figure S10.**
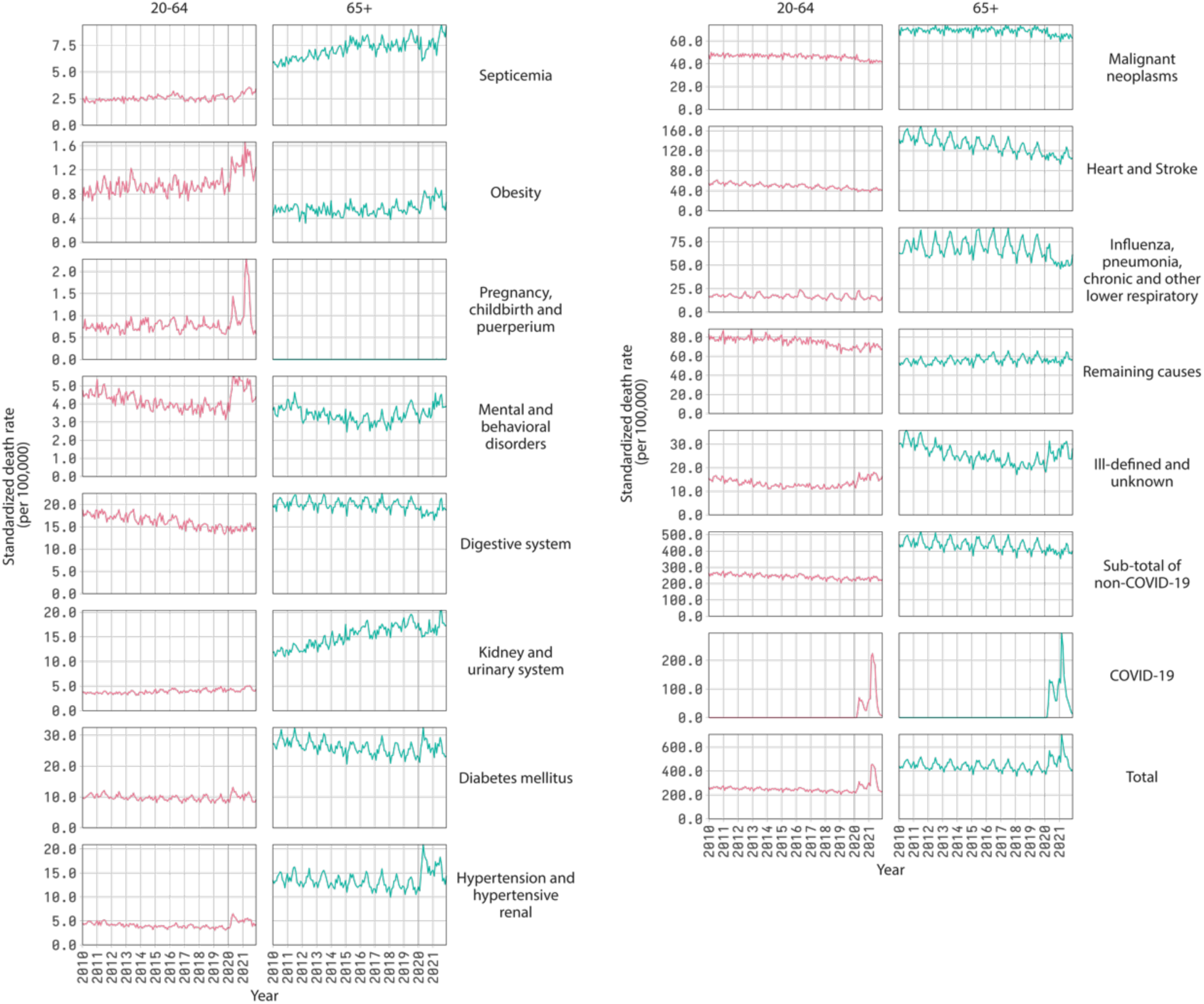
Time series of monthly standardized death rates in Brazil by groups of causes of death and major age groups 20-64 and 65+, Brazil, 2010-2011. The reader is cautioned that the vertical scales of the panels are different to uncover variations from each cause of death.

**Figure S11.**
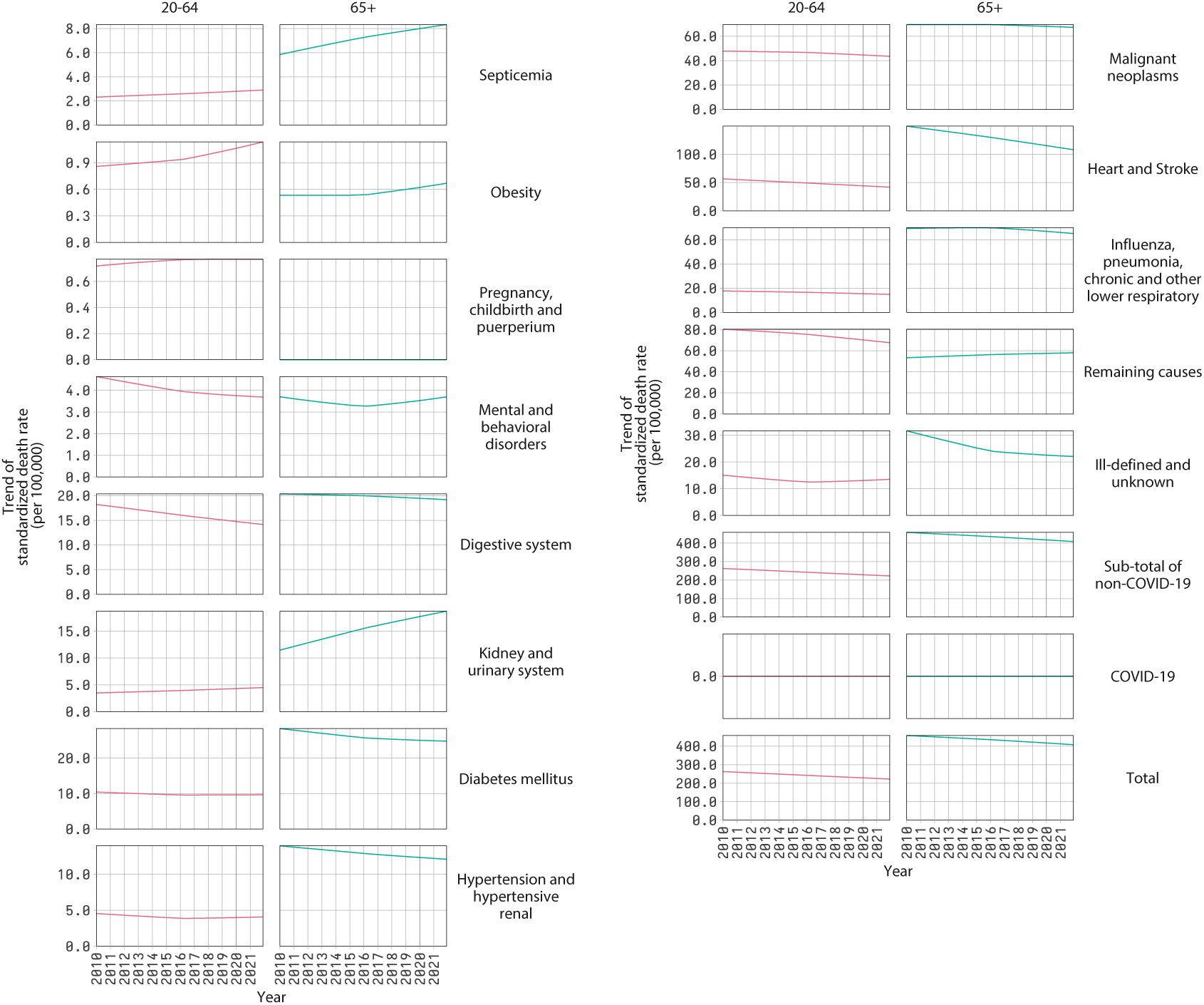
Trend component of the decomposition of time series of monthly standardized death rates by groups of cause of death and major age groups 20-64 and 65+, Brazil, 2020-2021. The graphs for “Total” are the sum of the trend component for each group of causes of death. The reader is cautioned that the vertical scales of the panels are different to uncover variations from each cause of death.

**Figure S12.**
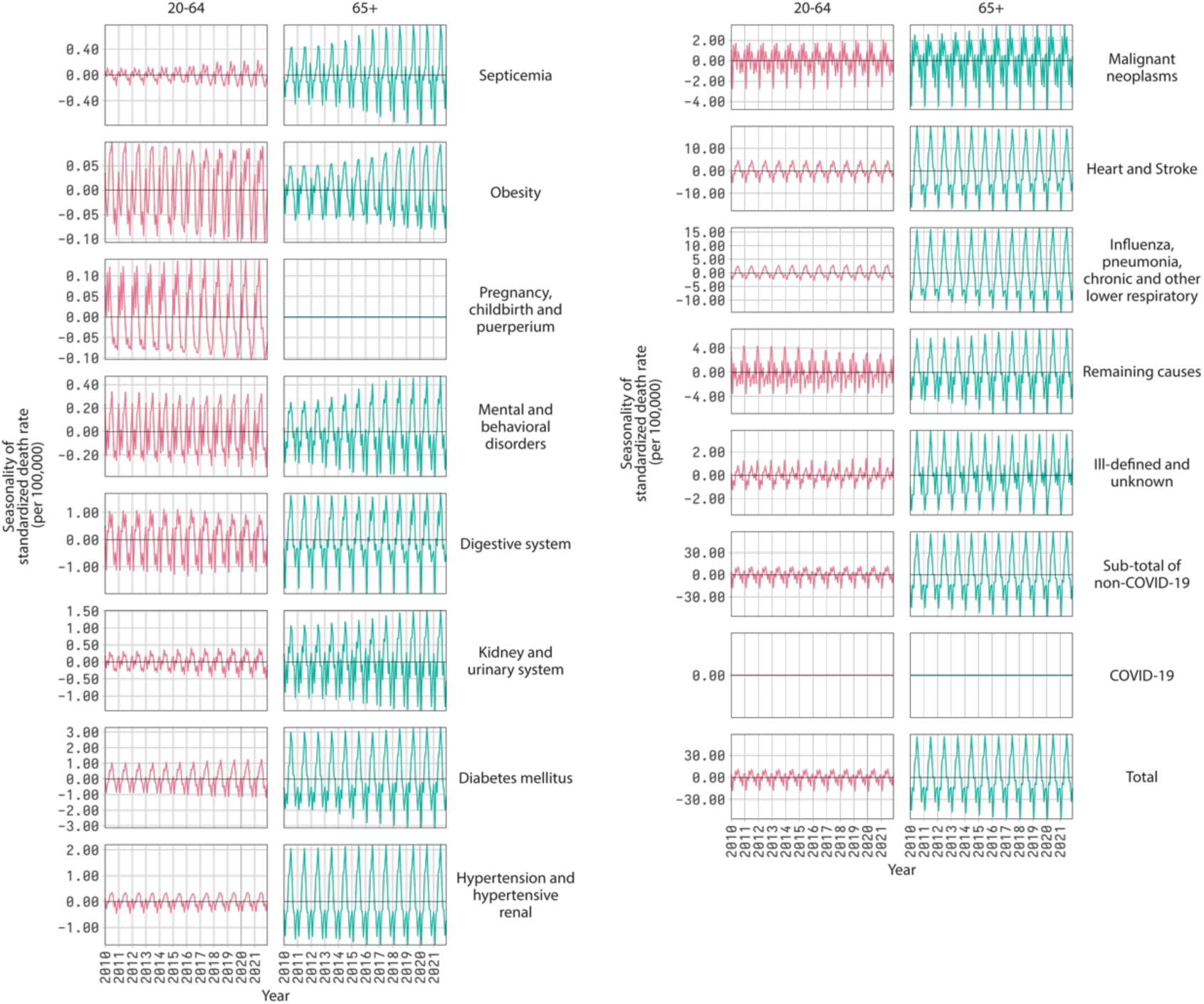
Seasonality component of the decomposition of time series of monthly standardized death rates by groups of cause of death and major age groups 20-64 and 65+, Brazil, 2020-2021. The graphs for “Total” are the sum of the seasonality component for each group of causes of death. The reader is cautioned that the vertical scales of the panels are different to uncover variations from each cause of death.

**Figure S13.**
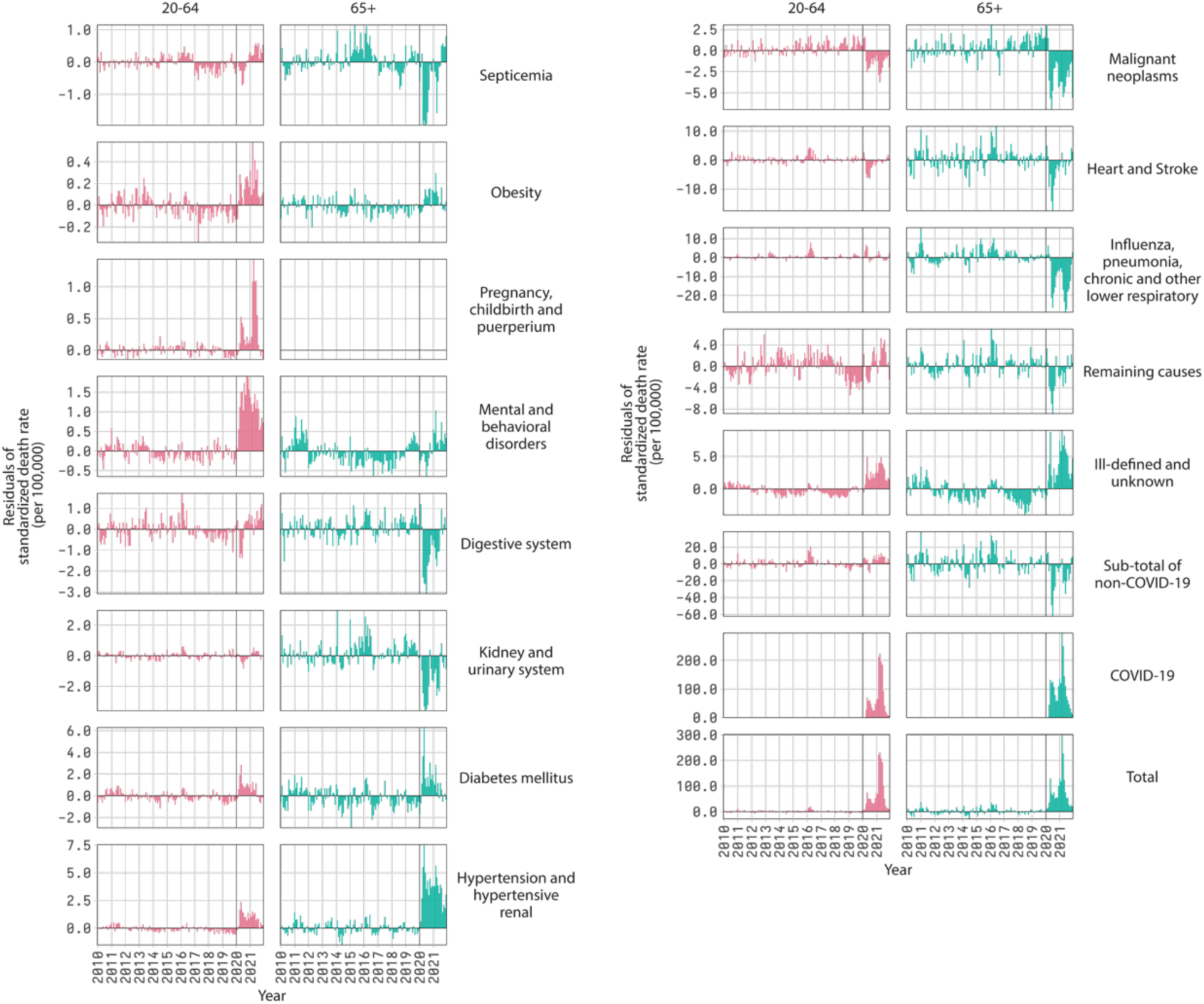
Remainder component of the decomposition of time series of monthly standardized death rates by groups of cause of death and major age groups 20-64 and 65+, Brazil, 2020-2021. The graphs for “Total” are the sum of the remainder component for each group of causes of death. The reader is cautioned that the vertical scales of the panels are different to uncover variations from each cause of death.

**Table S2.**
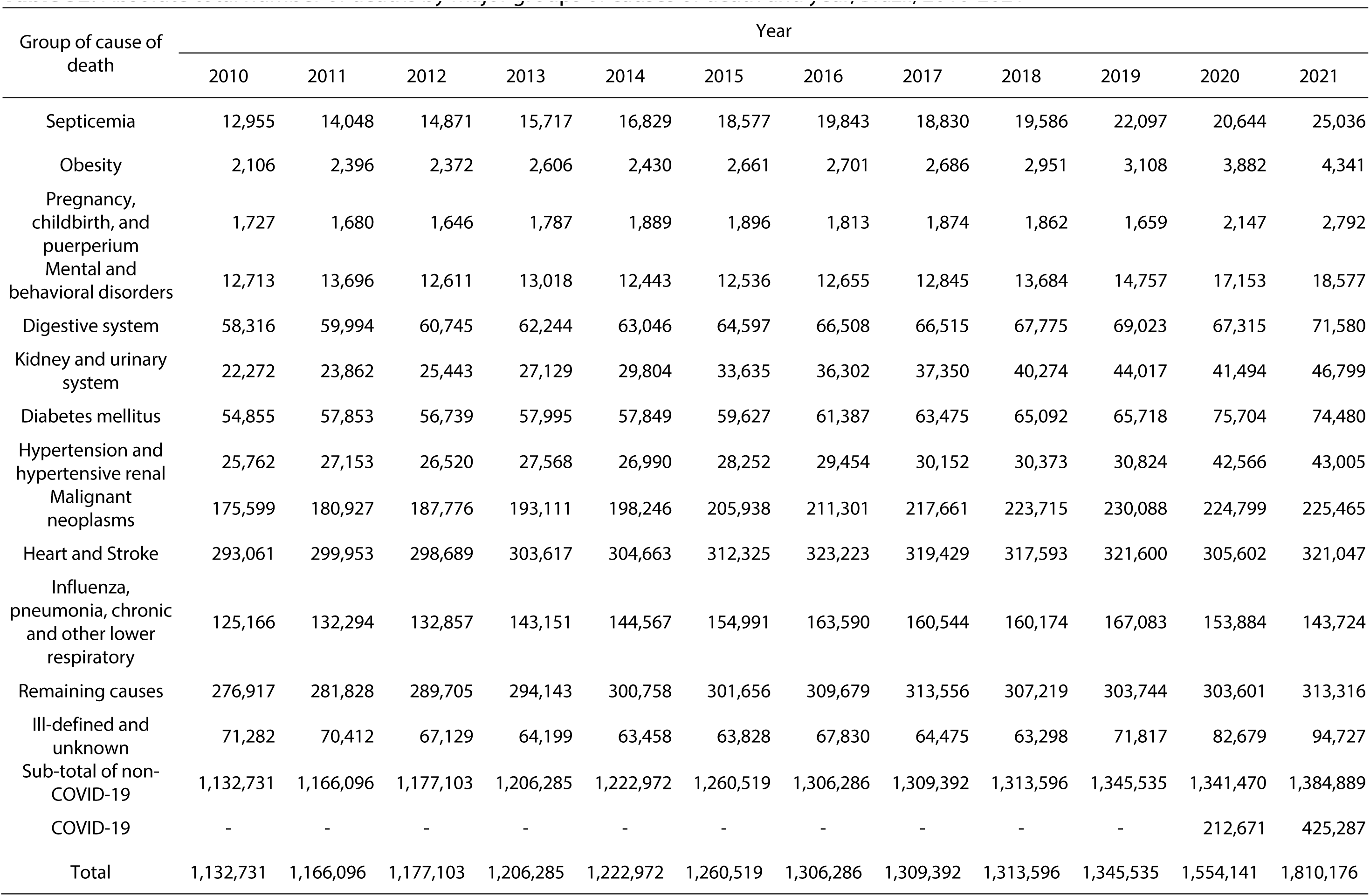
Absolute total number of deaths by major groups of causes of death and year, Brazil, 2010-2021

**Table S3.** Change in the absolute total number of deaths compared to previous year by major groups of causes of death and period, Brazil, 2010-2021

| Group of cause of death | Period |  |  |  |  |  |  |  |  |  |  |  |
| --- | --- | --- | --- | --- | --- | --- | --- | --- | --- | --- | --- | --- |
|  | 2011-2010 | 2012-2011 | 2013-2012 | 2014-2013 | 2015-2014 | 2016-2015 | 2017-2016 | 2018-2017 | 2019-2018 | 2020-2019 | 2021-2020 | Average<br>2017-2016 to<br>2019-2018 |
| Septicemia | 1,093 | 823 | 846 | 1,112 | 1,748 | 1,266 | -1,013 | 756 | 2,511 | -1,453 | 4,392 | 751 |
| Obesity | 290 | -24 | 234 | -176 | 231 | 40 | -15 | 265 | 157 | 774 | 459 | 136 |
| Pregnancy, childbirth,<br>and puerperium | -47 | -34 | 141 | 102 | 7 | -83 | 61 | -12 | -203 | 488 | 645 | -51 |
| Mental and behavioral<br>disorders | 983 | -1,085 | 407 | -575 | 93 | 119 | 190 | 839 | 1,073 | 2,396 | 1,424 | 701 |
| Digestive system | 1,678 | 751 | 1,499 | 802 | 1,551 | 1,911 | 7 | 1,260 | 1,248 | -1,708 | 4,265 | 838 |
| Kidney and urinary<br>system | 1,590 | 1,581 | 1,686 | 2,675 | 3,831 | 2,667 | 1,048 | 2,924 | 3,743 | -2,523 | 5,305 | 2,572 |
| Diabetes mellitus | 2,998 | -1,114 | 1,256 | -146 | 1,778 | 1,760 | 2,088 | 1,617 | 626 | 9,986 | -1,224 | 1,444 |
| Hypertension and<br>hypertensive renal | 1,391 | -633 | 1,048 | -578 | 1,262 | 1,202 | 698 | 221 | 451 | 11,742 | 439 | 457 |
| Malignant neoplasms | 5,328 | 6,849 | 5,335 | 5,135 | 7,692 | 5,363 | 6,360 | 6,054 | 6,373 | -5,289 | 666 | 6,262 |
| Heart and Stroke | 6,892 | -1,264 | 4,928 | 1,046 | 7,662 | 10,898 | -3,794 | -1,836 | 4,007 | -15,998 | 15,445 | -541 |
| Influenza, pneumonia,<br>chronic and other lower<br>respiratory | 7,128 | 563 | 10,294 | 1,416 | 10,424 | 8,599 | -3,046 | -370 | 6,909 | -13,199 | -10,160 | 1,164 |
| Remaining causes | 4,911 | 7,877 | 4,438 | 6,615 | 898 | 8,023 | 3,877 | -6,337 | -3,475 | -143 | 9,715 | -1,978 |
| Ill-defined and<br>unknown | -870 | -3,283 | -2,930 | -741 | 370 | 4,002 | -3,355 | -1,177 | 8,519 | 10,862 | 12,048 | 1,329 |
| Sub-total of non-COVID-19 | 33,365 | 11,007 | 29,182 | 16,687 | 37,547 | 45,767 | 3,106 | 4,204 | 31,939 | -4,065 | 43,419 | 33,365 |
| COVID-19 | - | - | - | - | - | - | - | - | - | - | 212,616 | - |
| Total | 33,365 | 11,007 | 29,182 | 16,687 | 37,547 | 45,767 | 3,106 | 4,204 | 31,939 | -4,065 | 256,035 | 13,083 |

**Table S4.** Absolute contribution of groups of causes of death to annual changes in the life expectancy at birth (e_0_) in Brazil, 2017-2020.

| Groups of causes of death | Period |  |  |  |
| --- | --- | --- | --- | --- |
|  | 2017-2018 | 2018-2019 | 2019-2020 | 2020-2021 |
| Septicemia | -0.001 | -0.017 | 0.020 | -0.030 |
| Obesity | -0.002 | -0.001 | -0.007 | -0.003 |
| Pregnancy, childbirth, and puerperium | 0.000 | 0.003 | -0.006 | -0.009 |
| Mental and behavioral disorders | -0.004 | -0.006 | -0.023 | -0.004 |
| Digestive system | 0.009 | 0.008 | 0.034 | -0.017 |
| Kidney and urinary system | -0.014 | -0.021 | 0.034 | -0.027 |
| Diabetes mellitus | 0.006 | 0.016 | -0.069 | 0.030 |
| Hypertension and hypertensive renal | 0.010 | 0.007 | -0.092 | 0.008 |
| Malignant neoplasms | 0.010 | 0.009 | 0.115 | 0.049 |
| Heart and Stroke | 0.130 | 0.070 | 0.240 | -0.040 |
| Influenza, pneumonia, chronic and other lower respiratory | 0.056 | -0.010 | 0.179 | 0.108 |
| Remaining causes | 0.179 | 0.154 | 0.091 | -0.004 |
| Ill-defined and unknown | 0.034 | -0.070 | -0.075 | -0.074 |
| Sub-total of non-COVID-19 | 0.413 | 0.142 | 0.440 | -0.013 |
| COVID-19 | 0.000 | 0.000 | -1.885 | -1.773 |
| Total | 0.413 | 0.142 | -1.444 | -1.785 |

**Table S5.**
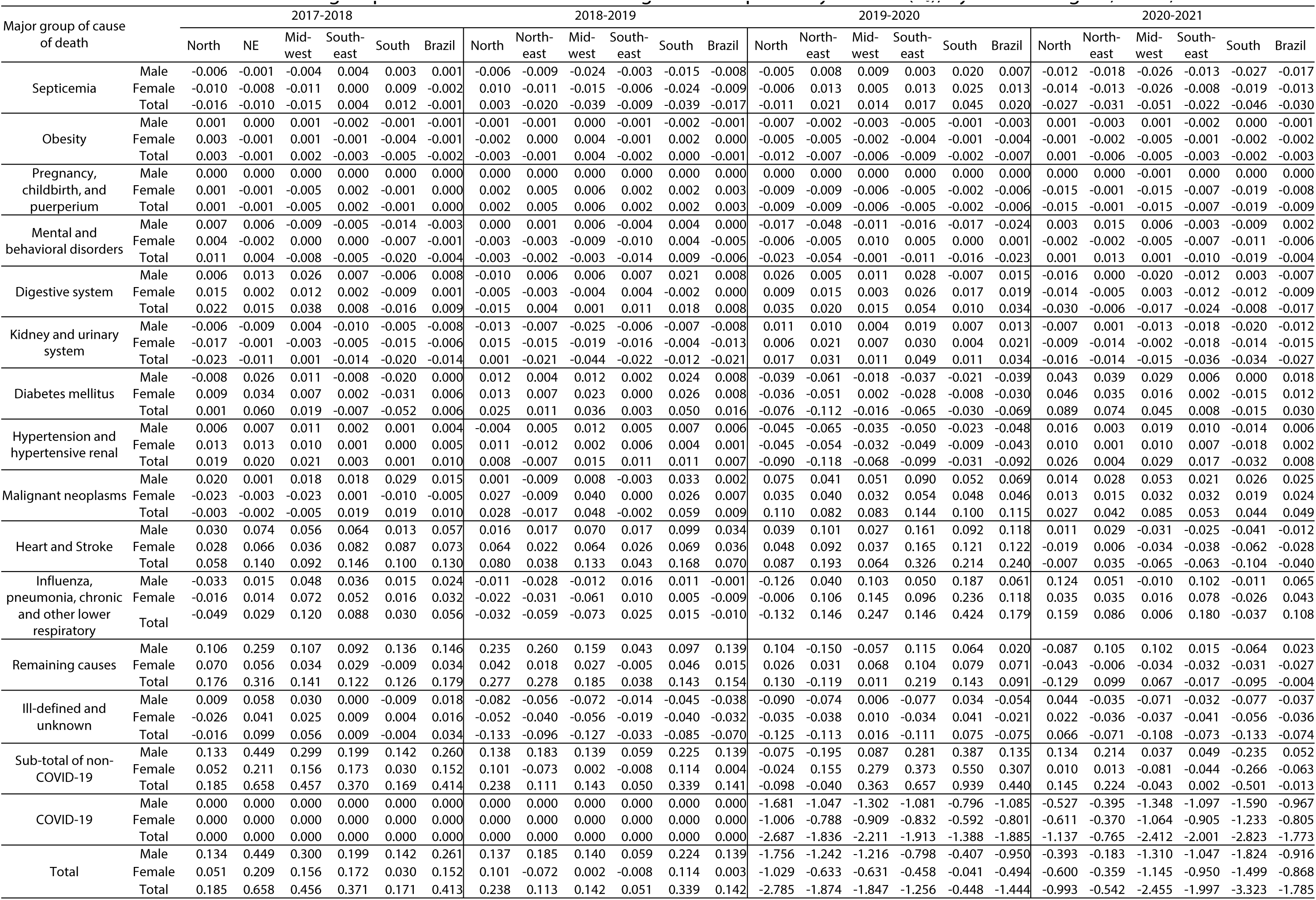
Absolute contribution of groups of causes of death to change in life expectancy at birth (e_0_), by sex and region, Brazil, 2017-2021

**Table S6.**
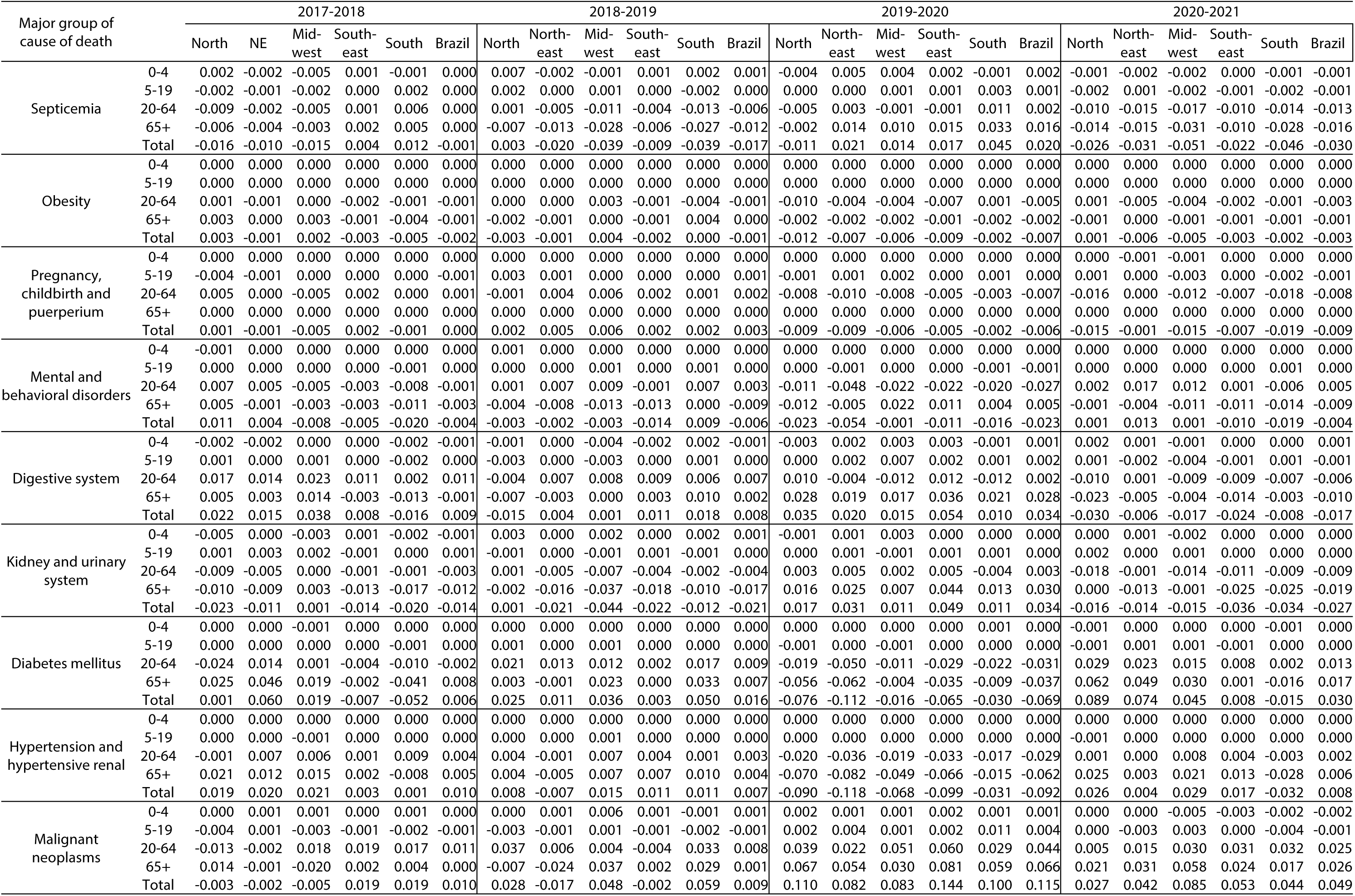
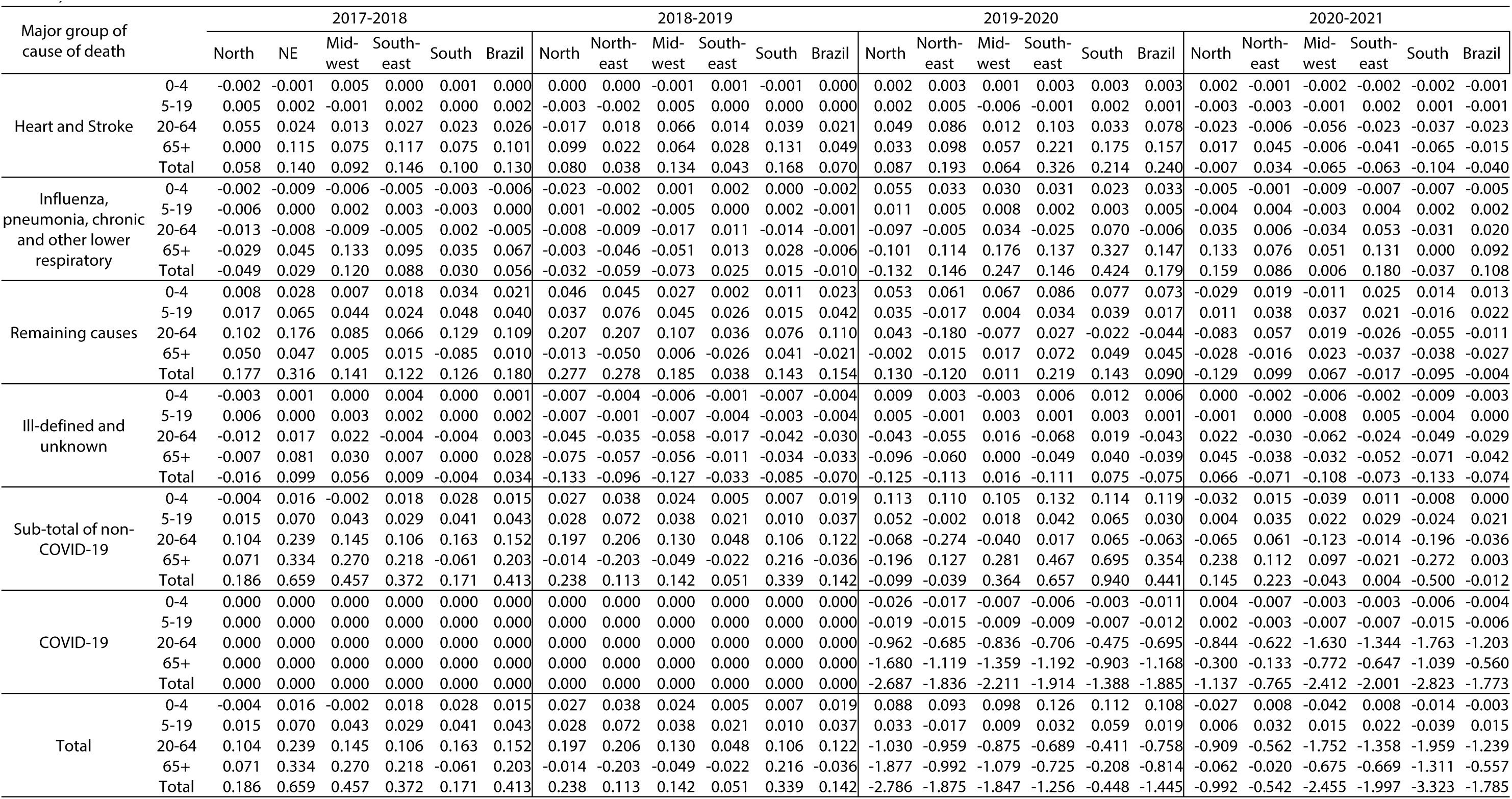
Absolute contribution of groups of causes of death to change in life expectancy at birth (e_0_), by major age groups and region, Brazil, 2017-2021

